# Projecting the transmission dynamics of SARS-CoV-2 through the post-pandemic period

**DOI:** 10.1101/2020.03.04.20031112

**Authors:** Stephen M. Kissler, Christine Tedijanto, Edward M. Goldstein, Yonatan H. Grad, Marc Lipsitch

## Abstract

There is an urgent need to project how transmission of the novel betacoronavirus SARS-CoV-2 will unfold in coming years. These dynamics will depend on seasonality, the duration of immunity, and the strength of cross-immunity to/from the other human coronaviruses. Using data from the United States, we measured how these factors affect transmission of human betacoronaviruses HCoV-OC43 and HCoV-HKU1. We then built a mathematical model to simulate transmission of SARS-CoV-2 through the year 2025. We project that recurrent wintertime outbreaks of SARS-CoV-2 will probably occur after an initial pandemic wave. We summarize the full range of plausible transmission scenarios and identify key data still needed to distinguish between them, most importantly longitudinal serological studies to determine the duration of immunity to SARS-CoV-2.

## Main text

The ongoing SARS-CoV-2 epidemic has caused nearly 80,000 detected cases of COVID-19 illness and claimed over 2,500 lives as of 24 Feb 2020 (*1*). With sustained transmission reported in China, Japan, Iran, Italy, and South Korea (*1*), the outbreak is on the verge of becoming a pandemic. The required intensity, duration, and urgency of public health responses will depend on how the initial pandemic wave unfolds and on the subsequent transmission dynamics of SARS-CoV-2. One possibility is that SARS-CoV-2 will follow its closest genetic relative, SARS-CoV, and be eradicated by intensive public health measures after causing a brief but intense epidemic (*2*). Increasingly, public health authorities consider this scenario unlikely (*3*). Alternatively, the transmission of SARS-CoV-2 could resemble that of pandemic influenza by circulating seasonally after causing an initial global wave of infection (*4*). Such a scenario could reflect the previous emergence of known human coronaviruses from zoonotic origins *e*.*g*. human coronavirus (HCoV) OC43 (*5*). This paper identifies viral, environmental, and immunologic factors which in combination will determine which scenarios in fact play out, and identifies key data needed to distinguish between them.

The transmission dynamics of the SARS-CoV-2 epidemic will depend on factors including the degree of seasonal variation in transmission strength, the duration of immunity, and the degree of cross-immunity between SARS-CoV-2 and other coronaviruses. SARS-CoV-2 belongs to the betacoronavirus genus, which includes the SARS coronavirus, MERS coronavirus, and two other human coronaviruses, HCoV-OC43 and HCoV-HKU1. The SARS and MERS coronaviruses cause severe illness with case fatality rates of 9 and 36% respectively, but the transmission of both has remained limited (*6*). HCoV-OC43 and HCoV-HKU1 infections may be asymptomatic or associated with mild to moderate upper respiratory tract illness; these HCoVs are considered the second most common cause of the common cold (*6*). While investigations into the spectrum of illness caused by SARS-CoV-2 are ongoing, recent evidence indicates the majority of cases experience mild illness with more limited occurrence of severe lower respiratory infection (*7*). Current COVID-19 case fatality rates in China are estimated at 2.9% within Hubei province, and 0.4% outside (*8*), suggesting lower severity than SARS and MERS but higher severity than HCoV-OC43 and HCoV-HKU1. In terms of transmission, the ability of SARS-CoV-2 to cause widespread infection is more reflective of HCoV-OC43 and HCoV-HKU1 than of its more clinically severe relatives. HCoV-OC43 and HCoV-HKU1 cause annual wintertime outbreaks of respiratory illness in temperate regions (*9, 10*), suggesting that wintertime climate and host behaviors may facilitate transmission, as is true for influenza (*11*–*13*). Immunity to HCoV-OC43 and HCoV-HKU1 appears to wane appreciably within one year (*14*), while SARS infection can induce longer-lasting immunity (*15*). The betacoronaviruses can induce immune responses against one another: SARS infection can generate neutralizing antibodies against HCoV-OC43 (*15*) and HCoV-OC43 infection can generate cross-reactive antibodies against SARS (*16*). However, due to the novelty of the SARS-CoV-2 epidemic and a relative lack of surveillance data on the existing human coronaviruses, it has not been possible to project how the transmission of SARS-CoV-2 will unfold in the coming years, including the interaction of SARS-CoV-2 with the seasonal coronaviruses.

We used data from the United States to model betacoronavirus transmission in temperate regions and to project the possible dynamics of SARS-CoV-2 infection through the year 2025. We first assessed the role of seasonal variation, duration of immunity, and cross immunity on the transmissibility of HCoV-OC43 and HCoV-HKU1 in the US. We used the weekly percentage of positive laboratory tests for HCoV-OC43 and HCoV-HKU1 (*17*) multiplied by the weekly population-weighted proportion of physician visits due to influenza-like illness (ILI) (*18, 19*) to approximate historical betacoronavirus incidence in the US to within a scaling constant. This proxy is proportional to incidence under a set of assumptions described in the Materials and Methods. To quantify variation in transmission strength over time, we estimated the weekly effective reproduction number, defined as the average number of secondary infections caused by a single infected individual (*20, 21*). The effective reproduction numbers for each of the betacoronaviruses displayed a seasonal pattern, with annual peaks in the effective reproduction number slightly preceding those of the incidence curves (**Fig S1**). For both HCoV-OC43 and HCoV-HKU1, the effective reproduction number typically reached its peak between October and November and its trough between February and April. Over the five seasons included in our data (2014-2019), the median effective reproduction number was 1.11 (IQR: 0.84-1.41) for HCoV-HKU1 and 1.00 (IQR: 0.84-1.41) for HCoV-OC43. Results were similar using various choices of incidence proxy and serial interval distributions (**Fig S1, Fig S2**).

To quantify the relative contribution of immunity *versus* seasonal forcing on the transmission dynamics of the betacoronaviruses, we adopted a regression model (*22*) that expressed the effective reproduction number for each strain (HKU1 and OC43) as the product of a baseline transmissibility constant (related to the basic reproduction number and the proportion of the population susceptible at the start of the season), the depletion of susceptibles due to infection with the same strain, the depletion of susceptibles due to infection with the other strain, and a spline to capture further unexplained seasonal variation in transmission strength (seasonal forcing). These covariates were able to explain most of the observed variability in the effective reproduction numbers (adjusted R^2^: 74.2%). The estimated multiplicative effects of each of these covariates on the weekly reproduction number are depicted in **Fig 1**. As expected, depletion of susceptibles for each strain was negatively correlated with transmissibility of that strain. Depletion of susceptibles for each strain was also negatively correlated with the reproduction number of the other betacoronavirus strain, providing evidence of cross-immunity. Per incidence proxy unit, the effect of the cross-immunizing strain was always less than the effect of the strain itself (**Table S1**), but the overall impact of cross-immunity could still be substantial if the cross-immunizing strain had a large outbreak (e.g. HCoV-OC43 in 2014-15 and 2016-17). Seasonal forcing appears to drive the rise in transmissibility at the start of the season (late October through early December), while depletion of susceptibles plays a comparatively larger role in the decline in transmissibility towards the end of the season. The strain-season coefficients were fairly consistent across seasons for each strain and lacked a clear correlation with incidence in prior seasons, consistent with experimental results showing substantial waning of immunity within a year (*14*).

**Figure 1.**
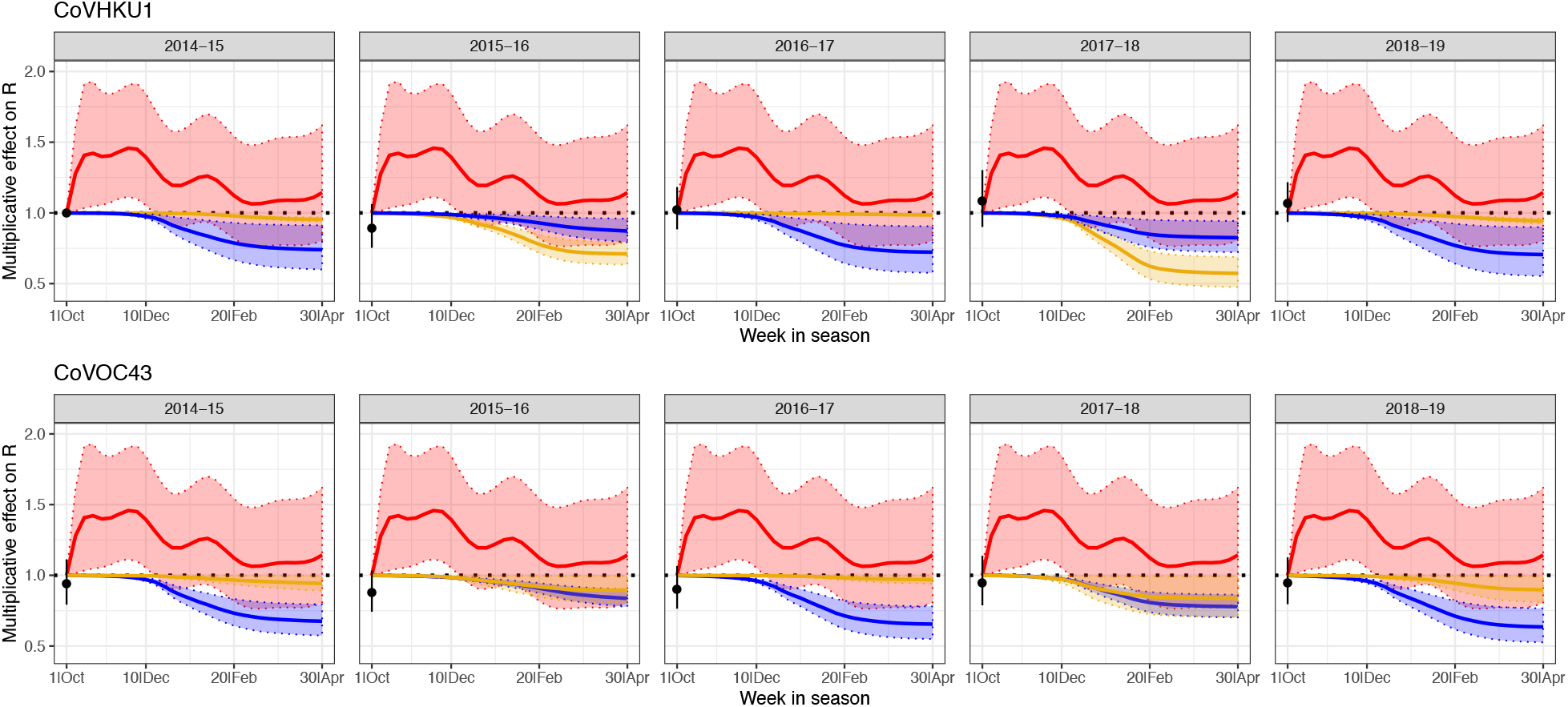
Estimated multiplicative effects of HCoV-HKU1 incidence (gold), HCoV-OC43 incidence (blue), and seasonal forcing (red) on weekly effective reproduction numbers of HCoV-HKU1 (top panels) and HCoV-OC43 (bottom), with 95% confidence intervals. The point at the start of each season is the estimated baseline for that strain and season compared to the 2014-15 HCoV-HKU1 season. The seasonal forcing spline is set to 1 at the first week of the season (no intercept).

We integrated these findings into a two-strain ordinary differential equation (ODE) susceptible-exposed-infectious-recovered (SEIR) compartmental model to describe the transmission dynamics of HCoV-OC43 and HCoV-HKU1 (**Fig S3**). The model provided a good fit to both the weekly incidence proxies for HCoV-OC43 and HCoV-HKU1 and to the estimated weekly effective reproduction numbers (**Fig 2**). According to the best-fit model parameters, the basic reproduction number for HCoV-OC43 and HCoV-HKU1 varies between 1.4 in the summer and 2 in the winter, the duration of immunity for both strains is about 40 weeks, and each strain induces cross-immunity against the other, though the cross-immunity that HCoV-OC43 infection induces against HCoV-HKU1 is stronger than the reverse.

**Figure 2.**
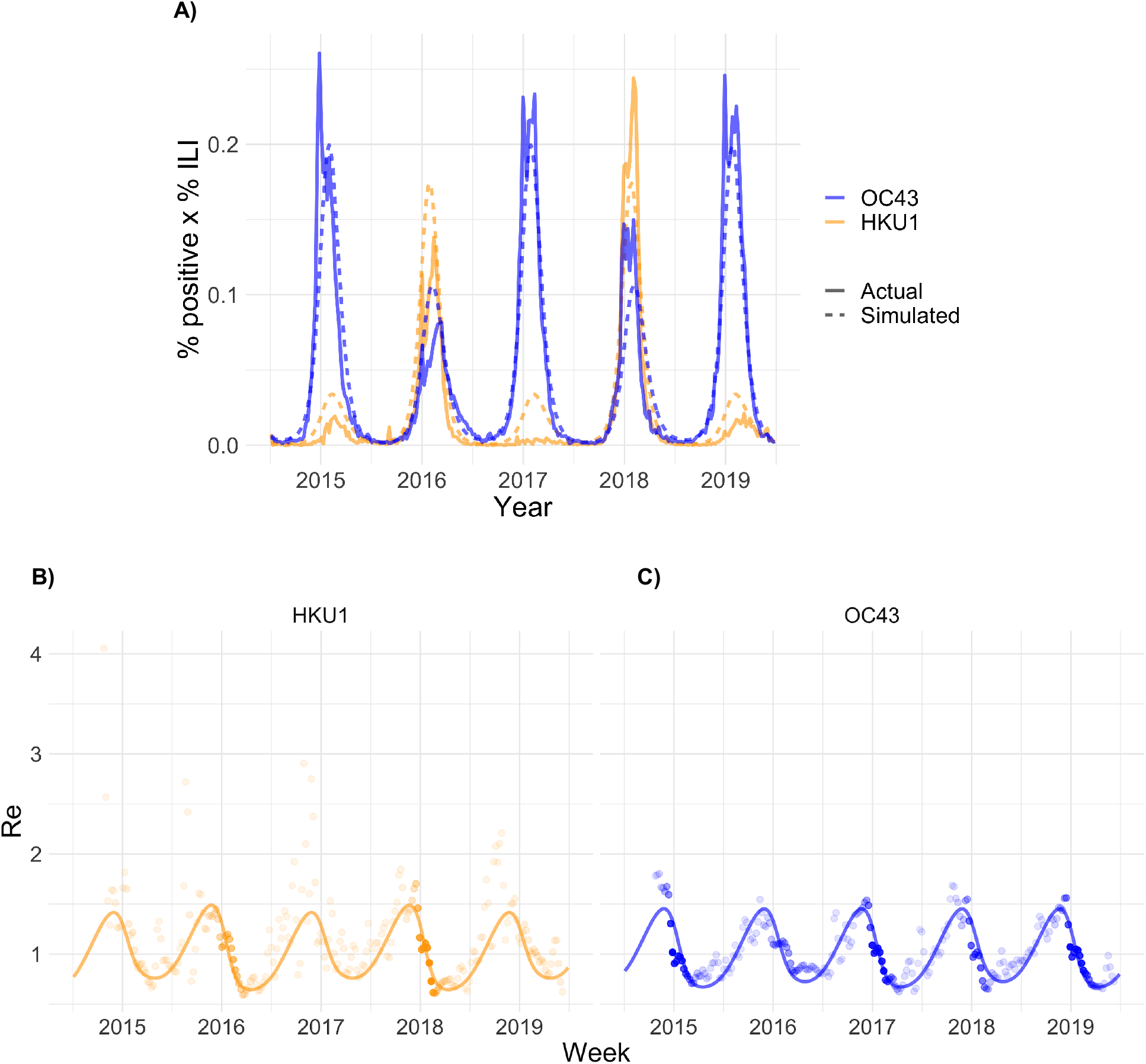
(A) Weekly percent positive laboratory tests x percent influenza-like illness (ILI) for the two common human betacoronaviruses in the United States between 5 July 2014 and 29 June 2019 (solid lines) with simulated output from the best-fit SEIR transmission model (dashed lines). (B-C) Weekly effective reproduction numbers (*R*_*e*_) estimated using the Wallinga-Teunis method (points) and simulated *R*_*e*_ from the best-fit SEIR transmission model (line) for the two common human betacoronaviruses. The opacity of each point is determined by the relative percent ILI x percent positive laboratory tests in that week relative to the maximum percent ILI x percent positive laboratory tests for that strain across the study period, which reflects uncertainty in the *R*_*e*_ estimate; estimates are more certain (darker points) in weeks with higher incidence.

Next, we incorporated a third betacoronavirus into the dynamic transmission model to represent SARS-CoV-2. We assumed that the incubation period and infectious period for SARS-CoV-2 were the same as the best-fit values for the other betacoronaviruses (5.0 and 4.9 days, respectively; see Table S7), in agreement with other estimates (*23*–*25*). We allowed the cross immunities, duration of immunity, degree of seasonal variation in transmissibility, and establishment time of sustained transmission to vary. For a representative set of parameter values within these ranges, we measured the annual incidence proxy due to SARS-CoV-2 (**Table 1, Tables S2-3**) and the annual SARS-CoV-2 outbreak peak size (**Tables S4-6**) for the five years following the simulated time of establishment. We summarized the post-pandemic SARS-CoV-2 dynamics into the categories of annual outbreaks, biennial outbreaks, sporadic outbreaks, or virtual elimination. Overall, shorter durations of immunity and smaller degrees of cross immunity from the other betacoronaviruses were associated with greater total incidence of infection due to SARS-CoV-2, and autumn establishments and smaller seasonal fluctuations in transmissibility were associated with larger pandemic peak sizes. Model simulations demonstrated the following few key points:

i. *SARS-CoV-2 can proliferate at any time of year*. In all modeled scenarios, SARS-CoV-2 was capable of producing a substantial outbreak regardless of establishment time. Winter/spring establishments favored longer-lasting outbreaks with shorter peaks (**Fig 3A**), while autumn/winter establishments led to more acute outbreaks (**Fig 3B**). The five-year cumulative incidence proxies were comparable for all establishment times (**Table 1**).
ii. *If immunity to SARS-CoV-2 is not permanent, it will likely enter into regular circulation*. Much like pandemic influenza, many scenarios lead to SARS-CoV-2 entering into long-term circulation alongside the other human betacoronaviruses (*e*.*g*. **Fig 3A–B**), possibly in annual, biennial, or sporadic patterns over the next five years (**Table 1**). Short-term immunity (on the order of 40 weeks, similar to HCoV-OC43 and HCoV-HKU1) favors the establishment of annual SARS-CoV-2 outbreaks, while longer-term immunity (two years) favors biennial outbreaks if establishment occurs in the winter or spring and sporadic outbreaks if establishment occurs in the summer or autumn.
iii. *If immunity to SARS-CoV-2 is permanent, the virus could disappear for five or more years after causing a major outbreak*. Long-term immunity consistently led to effective elimination of SARS-CoV-2 and lower overall incidence of infection. If SARS-CoV-2 induces cross immunity against HCoV-OC43 and HCoV-HKU1, the incidence of all betacoronaviruses could decline and even virtually disappear (**Fig 3C**). The virtual elimination of HCoV-OC43 and HCoV-HKU1 would be possible if SARS-CoV-2 induced 70% cross immunity against them, which is the same estimated level of cross-immunity that HCoV-OC43 induces against HCoV-HKU1.
iv. *Low levels of cross immunity from the other betacoronaviruses against SARS-CoV-2 could make SARS-CoV-2 appear to die out, only to resurge after a few years*. Even if SARS-CoV-2 immunity only lasts for two years, mild (30%) cross-immunity from HCoV-OC43 and HCoV-HKU1 could effectively eliminate the transmission of SARS-CoV-2 for up to three years before a resurgence in 2025, as long as SARS-CoV-2 does not fully die out (**Fig 3D**).
v. *The dynamics of coronavirus outbreaks in temperate regions over the next five years may depend heavily on the timing of SARS-CoV-2 establishment*. Under certain scenarios, altering just the timing of SARS-CoV-2 establishment made the difference between annual short-peaked outbreaks and more sporadic acute outbreaks in the post-pandemic period (**Fig 3E–F)**. The establishment of sustained transmission can be delayed by rapidly detecting and isolating introduced cases (*1*).

**Table 1.** Cumulative projected percent influenza-like illness (ILI) x percent positive laboratory tests for SARS-CoV-2 by year for a representative set of cross immunities, immunity durations, and establishment times. Percent ILI is measured as the population-weighted proportion of visits to sentinel providers that were associated with influenza symptoms. The seasonality factor represents the amount of seasonal variation in the SARS-CoV-2 *R*_*0*_ relative to the other human betacoronaviruses. A seasonality factor of 1 indicates equivalent seasonal variation in *R*_*0*_, while 0 indicates no seasonal variation (see **Table S2-3**). Chi3X represents the degree of cross-immunity induced by infection with SARS-CoV-2 against OC43 and HKU1 and ChiX3 represents the degree of cross-immunity induced by OC43 or HKU1 infection against SARS-CoV-2. The establishment times correspond to: Winter - week 4 (early February); Spring - week 16 (late April); Summer - week 28 (mid July); Autumn - week 40 (early October). Darker shading corresponds to higher cumulative infection sizes.

| Chi3X | ChiX3 | Immunity (wks) | Est. time | Seasonality factor | 2019 | 2020 | 2021 | 2022 | 2023 | 2024 | 2025 | Max | Post-pandemic transmission |
| --- | --- | --- | --- | --- | --- | --- | --- | --- | --- | --- | --- | --- | --- |
| 0.7 | 0 | 40 | Winter | 1 | 0.00 | 5.04 | 3.55 | 4.18 | 3.98 | 3.96 | 3.99 | 24.70 | Annual |
| 0.7 | 0.3 | 40 | Winter | 1 | 0.00 | 2.44 | 4.64 | 4.06 | 2.57 | 4.26 | 2.82 | 20.78 | Annual |
| 0.3 | 0 | 40 | Winter | 1 | 0.00 | 5.04 | 3.55 | 4.18 | 3.98 | 3.96 | 3.99 | 24.70 | Annual |
| 0 | 0 | 40 | Winter | 1 | 0.00 | 5.04 | 3.55 | 4.18 | 3.98 | 3.96 | 3.99 | 24.70 | Annual |
| 0.7 | 0 | 104 | Winter | 1 | 0.00 | 4.79 | 0.17 | 2.60 | 0.33 | 3.17 | 0.02 | 11.08 | Biennial |
| 0.7 | 0.3 | 104 | Winter | 1 | 0.00 | 2.35 | 2.95 | 0.18 | 2.20 | 0.62 | 2.43 | 10.72 | Biennial |
| 0.3 | 0 | 104 | Winter | 1 | 0.00 | 4.79 | 0.17 | 2.60 | 0.33 | 3.17 | 0.02 | 11.08 | Biennial |
| 0 | 0 | 104 | Winter | 1 | 0.00 | 4.79 | 0.17 | 2.60 | 0.33 | 3.17 | 0.02 | 11.08 | Biennial |
| 0.7 | 0 | Permanent | Winter | 1 | 0.00 | 4.64 | 0.04 | 0.00 | 0.00 | 0.00 | 0.00 | 4.68 | Virtual elimination |
| 0.7 | 0.3 | Permanent | Winter | 1 | 0.00 | 2.29 | 1.62 | 0.00 | 0.00 | 0.00 | 0.00 | 3.91 | Virtual elimination |
| 0.3 | 0 | Permanent | Winter | 1 | 0.00 | 4.64 | 0.04 | 0.00 | 0.00 | 0.00 | 0.00 | 4.68 | Virtual elimination |
| 0 | 0 | Permanent | Winter | 1 | 0.00 | 4.64 | 0.04 | 0.00 | 0.00 | 0.00 | 0.00 | 4.68 | Virtual elimination |
| 0.7 | 0 | 40 | Spring | 1 | 0.00 | 1.03 | 5.94 | 4.67 | 3.97 | 3.94 | 4.01 | 23.56 | Annual |
| 0.7 | 0.3 | 40 | Spring | 1 | 0.00 | 0.36 | 5.89 | 4.04 | 2.88 | 3.99 | 2.84 | 19.99 | Annual |
| 0.3 | 0 | 40 | Spring | 1 | 0.00 | 1.03 | 5.94 | 4.67 | 3.97 | 3.94 | 4.01 | 23.56 | Annual |
| 0 | 0 | 40 | Spring | 1 | 0.00 | 1.03 | 5.94 | 4.67 | 3.97 | 3.94 | 4.01 | 23.56 | Annual |
| 0.7 | 0 | 104 | Spring | 1 | 0.00 | 1.02 | 4.53 | 0.04 | 3.61 | 0.01 | 2.11 | 11.32 | Biennial |
| 0.7 | 0.3 | 104 | Spring | 1 | 0.00 | 0.36 | 5.04 | 0.00 | 0.52 | 3.69 | 0.01 | 9.61 | Sporadic |
| 0.3 | 0 | 104 | Spring | 1 | 0.00 | 1.02 | 4.53 | 0.04 | 3.61 | 0.01 | 2.10 | 11.32 | Biennial |
| 0 | 0 | 104 | Spring | 1 | 0.00 | 1.02 | 4.53 | 0.04 | 3.61 | 0.01 | 2.10 | 11.32 | Biennial |
| 0.7 | 0 | Permanent | Spring | 1 | 0.00 | 1.02 | 3.99 | 0.00 | 0.00 | 0.00 | 0.00 | 5.00 | Virtual elimination |
| 0.7 | 0.3 | Permanent | Spring | 1 | 0.00 | 0.36 | 4.58 | 0.00 | 0.00 | 0.00 | 0.00 | 4.94 | Virtual elimination |
| 0.3 | 0 | Permanent | Spring | 1 | 0.00 | 1.02 | 3.99 | 0.00 | 0.00 | 0.00 | 0.00 | 5.00 | Virtual elimination |
| 0 | 0 | Permanent | Spring | 1 | 0.00 | 1.02 | 3.99 | 0.00 | 0.00 | 0.00 | 0.00 | 5.00 | Virtual elimination |
| 0.7 | 0 | 40 | Summer | 1 | 0.00 | 0.00 | 6.64 | 4.45 | 3.80 | 4.00 | 4.00 | 22.90 | Annual |
| 0.7 | 0.3 | 40 | Summer | 1 | 0.00 | 0.00 | 6.15 | 3.51 | 3.58 | 3.13 | 3.88 | 20.25 | Annual |
| 0.3 | 0 | 40 | Summer | 1 | 0.00 | 0.00 | 6.64 | 4.45 | 3.80 | 4.00 | 4.00 | 22.90 | Annual |
| 0 | 0 | 40 | Summer | 1 | 0.00 | 0.00 | 6.64 | 4.45 | 3.80 | 4.00 | 4.00 | 22.90 | Annual |
| 0.7 | 0 | 104 | Summer | 1 | 0.00 | 0.00 | 6.05 | 0.00 | 0.09 | 4.41 | 0.00 | 10.56 | Sporadic |
| 0.7 | 0.3 | 104 | Summer | 1 | 0.00 | 0.00 | 5.58 | 0.00 | 0.03 | 3.89 | 0.01 | 9.51 | Sporadic |
| 0.3 | 0 | 104 | Summer | 1 | 0.00 | 0.00 | 6.05 | 0.00 | 0.10 | 4.39 | 0.00 | 10.55 | Sporadic |
| 0 | 0 | 104 | Summer | 1 | 0.00 | 0.00 | 6.05 | 0.00 | 0.10 | 4.39 | 0.00 | 10.55 | Sporadic |
| 0.7 | 0 | Permanent | Summer | 1 | 0.00 | 0.00 | 5.77 | 0.00 | 0.00 | 0.00 | 0.00 | 5.77 | Virtual elimination |
| 0.7 | 0.3 | Permanent | Summer | 1 | 0.00 | 0.00 | 5.28 | 0.00 | 0.00 | 0.00 | 0.00 | 5.28 | Virtual elimination |
| 0.3 | 0 | Permanent | Summer | 1 | 0.00 | 0.00 | 5.77 | 0.00 | 0.00 | 0.00 | 0.00 | 5.77 | Virtual elimination |
| 0 | 0 | Permanent | Summer | 1 | 0.00 | 0.00 | 5.77 | 0.00 | 0.00 | 0.00 | 0.00 | 5.77 | Virtual elimination |
| 0.7 | 0 | 40 | Autumn | 1 | 0.00 | 0.00 | 6.61 | 3.20 | 4.15 | 4.16 | 3.93 | 22.05 | Annual |
| 0.7 | 0.3 | 40 | Autumn | 1 | 0.00 | 0.00 | 5.95 | 2.46 | 4.08 | 2.99 | 4.02 | 19.51 | Annual |
| 0.3 | 0 | 40 | Autumn | 1 | 0.00 | 0.00 | 6.61 | 3.20 | 4.15 | 4.16 | 3.93 | 22.05 | Annual |
| 0 | 0 | 40 | Autumn | 1 | 0.00 | 0.00 | 6.61 | 3.20 | 4.15 | 4.16 | 3.93 | 22.05 | Annual |
| 0.7 | 0 | 104 | Autumn | 1 | 0.00 | 0.00 | 6.25 | 0.00 | 0.00 | 3.75 | 0.04 | 10.04 | Sporadic |
| 0.7 | 0.3 | 104 | Autumn | 1 | 0.00 | 0.00 | 5.60 | 0.00 | 0.00 | 2.71 | 0.79 | 9.10 | Sporadic |
| 0.3 | 0 | 104 | Autumn | 1 | 0.00 | 0.00 | 6.25 | 0.00 | 0.00 | 3.73 | 0.05 | 10.03 | Sporadic |
| 0 | 0 | 104 | Autumn | 1 | 0.00 | 0.00 | 6.25 | 0.00 | 0.00 | 3.73 | 0.05 | 10.03 | Sporadic |
| 0.7 | 0 | Permanent | Autumn | 1 | 0.00 | 0.00 | 6.06 | 0.00 | 0.00 | 0.00 | 0.00 | 6.06 | Virtual elimination |
| 0.7 | 0.3 | Permanent | Autumn | 1 | 0.00 | 0.00 | 5.40 | 0.00 | 0.00 | 0.00 | 0.00 | 5.40 | Virtual elimination |
| 0.3 | 0 | Permanent | Autumn | 1 | 0.00 | 0.00 | 6.06 | 0.00 | 0.00 | 0.00 | 0.00 | 6.06 | Virtual elimination |
| 0 | 0 | Permanent | Autumn | 1 | 0.00 | 0.00 | 6.06 | 0.00 | 0.00 | 0.00 | 0.00 | 6.06 | Virtual elimination |

**Figure 3.**
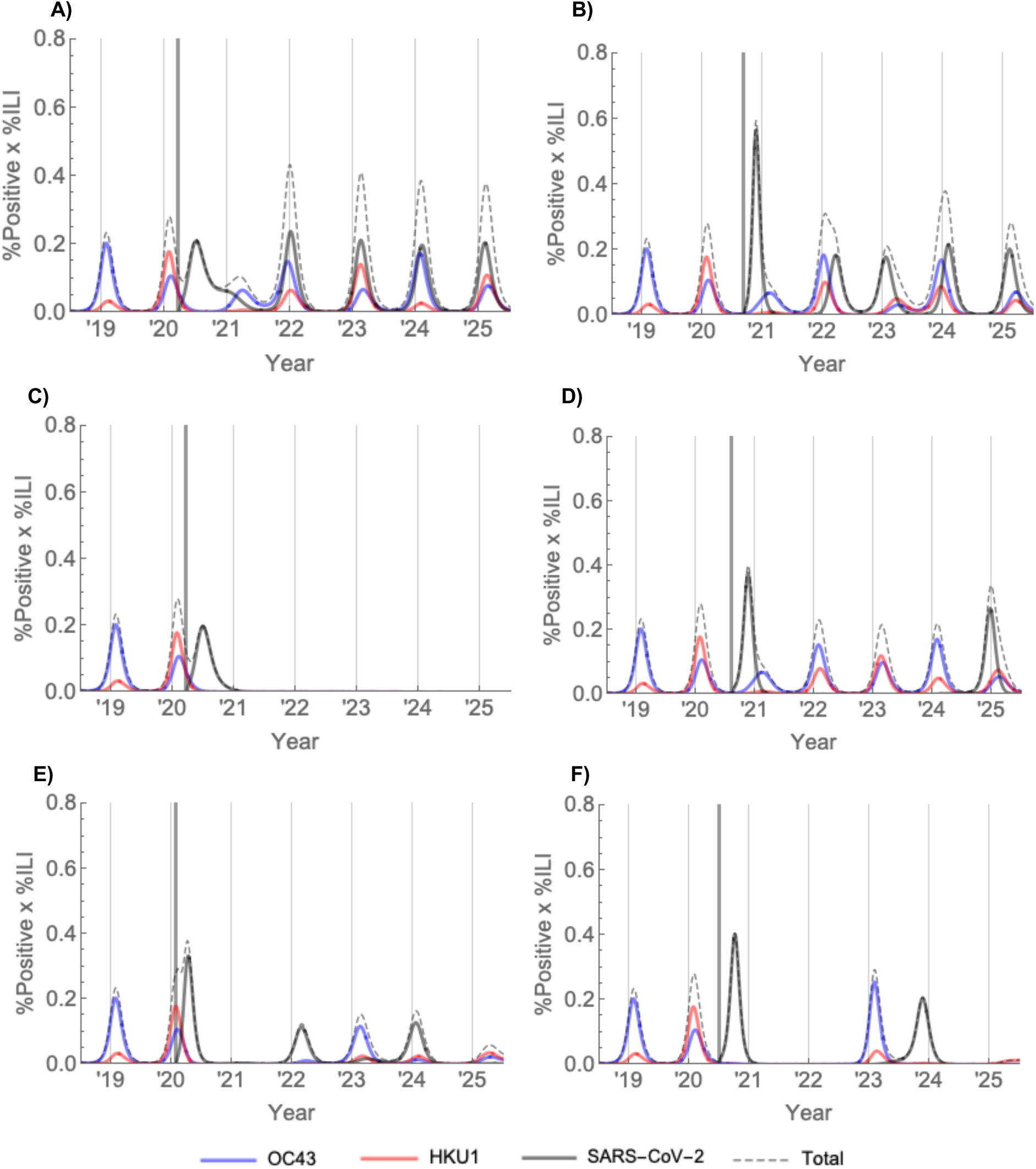
A representative set of invasion scenarios for SARS-CoV-2 in the US. The thick vertical bar marks the establishment time of SARS-CoV-2. *(A) χ*_*3X*_ = 0.3, *χ*_*X3*_ = 0, *σ*_*3*_ = 1/40, *t*_*est*_ = week 12 (late March) *(B) χ*_*3X*_ = 0.3, *χ*_*X3*_ = 0, *σ*_*3*_ = 1/40, *t*_*est*_ = week 36 (early September) *(C) χ*_*3X*_ = 0.7, *χ*_*X3*_ = 0, *σ*_*3*_ = 0, *t*_*est*_ = week 12 (late March) (D) *χ*_*3X*_ = 0.3, *χ*_*X3*_ = 0.3, *σ*_*3*_ = 1/104, *t*_*est*_ = week 32 (mid August) *(E) χ*_*3X*_ = 0.7, *χ*_*X3*_ = 0, *σ*_*3*_ = 1/104, *t*_*est*_ = week 4 (early February) *(F) χ*_*3X*_ = 0.7, *χ*_*X3*_ = 0, *σ*_*3*_ = 1/104, *t*_*est*_ = week 26 (late June)

Our observations are consistent with other predictions of how the SARS-CoV-2 outbreak might unfold. According to a study of geographic variation in the SARS-CoV-2 basic reproduction number across China, seasonal variations in absolute humidity will be insufficient to prevent the widespread transmission of SARS-CoV-2 (*26*). A modelling study using data from Sweden also found that seasonal establishment of SARS-CoV-2 transmission is likely in the post-pandemic period (*10*).

Our study was subject to a variety of limitations. Only five seasons of observational data on coronaviruses were available, though the incidence patterns resemble those from 10 years of data from a hospital in Sweden (*10*). We assumed that the spline coefficients were constant across all seasons though seasonal forcing likely differed from year to year based on underlying drivers. To keep the transmission model from becoming unreasonably complex, we assumed that there was no difference in the seasonal forcing, per-case force of infection, incubation period, or infectious period across betacoronaviruses. However, our estimates for these values lie within the ranges of estimates from the literature, so we do not anticipate that this detracted from our results. We also did not directly model any effect from the opening of schools, which could lead to an additional boost in transmission strength in the early autumn (*27*), or any effects of behavior change or control efforts, which could suppress the effective reproduction number. The transmission model is deterministic, so it cannot capture the possibility of SARS-CoV-2 extinction.

Accurately quantifying the probability of SARS-CoV-2 extinction would depend on many factors for which sufficient evidence is currently lacking. We have used percent test-positive multiplied by percent ILI to approximate coronavirus incidence up to a proportional constant; results were similar when using the raw number of positive tests and the raw percent positive as incidence proxies, see **Figure S1**. While the percent test-positive multiplied by percent ILI has been shown to be one of the best available proxies for influenza incidence (*18*), the conversion between this measure and true incidence of coronavirus infections is unclear, and so we do not make precise estimates of the overall coronavirus incidence. This conversion will undoubtedly depend on the particular population for which these estimates are being made. Recent evidence from New York suggests that some 4% of coronavirus cases seek medical care, and only a fraction of these are tested (*28*). In addition, the method that we adopted to estimate the effective reproduction number depends on the serial interval distribution, which has not been well-studied for commonly circulating human coronaviruses; we used the best-available evidence from SARS, the most closely related coronavirus to SARS-CoV-2. Our findings only generalize to temperate regions, comprising 60% of the world’s population (*29*), and differences between average interpersonal contact rates between countries could further modulate the size and intensity of outbreaks. The transmission dynamics of respiratory illnesses in tropical regions can be much more complex. However, we expect that if post-pandemic transmission of SARS-CoV-2 does take hold in temperate regions, there will also be continued transmission in tropical regions seeded by the seasonal outbreaks to the north and south. With such reseeding, long-term disappearance of any strain becomes less likely (*30*), but according to our model the effective reproductive number of SARS-CoV-2 remains below 1 during most of each period when that strain disappears, meaning that reseeding would shorten these disappearances only modestly.

Our findings indicate key pieces of information that are still required to know how the current SARS-CoV-2 outbreak will unfold. Most crucially, longitudinal serological studies from patients infected with SARS-CoV-2 could indicate whether or not immunity wanes, and at what rate. According to our projections, this rate is the key modulator of the total SARS-CoV-2 incidence in the coming years. While long-lasting immunity would lead to lower overall incidence of infection, it would also complicate vaccine efficacy trials by contributing to low case numbers when those trials are conducted, as occurred with Zika virus (*31*). Furthermore, our findings underscore the need to maintain SARS-CoV-2 surveillance even if the outbreak appears to die out after the first pandemic wave, as a resurgence in infection could be possible as late as 2025.

In summary, the total incidence of COVID-19 illness over the next five years will depend critically upon whether or not it enters into regular circulation after the initial pandemic wave, which in turn depends primarily upon the duration of immunity that SARS-CoV-2 infection imparts. The intensity and timing of pandemic and post-pandemic outbreaks will depend on the time of year when widespread SARS-CoV-2 infection becomes established and, to a lesser degree, upon the magnitude of seasonal variation in transmissibility and the level of cross-immunity that exists between the betacoronaviruses. Longitudinal serological studies are urgently required to determine the duration of immunity to SARS-CoV-2, and epidemiological surveillance should be maintained in the coming years to anticipate the possibility of resurgence.

## Data Availability

A data use agreement with the CDC is required to access the NREVSS human coronavirus dataset. ILINet data is publicly available through the FluView Interactive website (19). Regression model code is available at https://github.com/ctedijanto/coronavirus-seasonality. Transmission model code is available at https://github.com/skissler/nCoV_introduction.

https://github.com/skissler/nCoV_introduction

## Acknowledgements

We thank Marie Killerby and Amber Haynes for their helpful comments on early versions of this manuscript.

## Author contributions

SMK conceived of the study, conducted the analysis, and wrote the manuscript. CT conceived of the study, conducted the analysis, and wrote the manuscript. E.M.G. assisted with the analysis and edited the manuscript. YHG edited the manuscript and oversaw the work. ML edited the manuscript and oversaw the work.

## Data and materials availability

A data use agreement with the CDC is required to access the NREVSS human coronavirus dataset. ILINet data is publicly available through the FluView Interactive website (*19*). Regression model code is available at https://github.com/ctedijanto/coronavirus-seasonality. Transmission model code is available at https://github.com/skissler/nCoV_introduction.

## Supplement

### Materials and Methods

#### Data

Viral testing data came from the US National Respiratory and Enteric Virus Surveillance System (NREVSS) (*17*). We extracted the weekly number of total tests for any coronavirus and positive tests for betacoronaviruses HCoV-OC43 and HCoV-HKU1 from all reporting laboratories between 5^th^ July 2014 and 29^th^ June 2019. Dividing the weekly number of positive tests by the total weekly number of tests yielded the weekly percentage of positive tests for each coronavirus.

To estimate the incidence of each betacoronavirus, we used an incidence proxy calculated by multiplying the weekly percentage of positive tests for each coronavirus by the weekly population-weighted proportion of physician visits due to influenza-like illness (ILI) (*13*). The assumptions needed for this proxy to capture true influenza incidence up to a multiplicative constant are described in Goldstein et al. (2011) (*18*). Since betacoronaviruses HCoV-OC43 and HCoV-HKU1 are more likely to cause milder cold-like symptoms or acute respiratory infection than ILI, our proxy additionally requires that the proportion of coronavirus cases with ILI is constant over the study period. The weekly proportions of ILI visits were obtained from the US Outpatient Influenza-like Illness Surveillance Network (ILINet) and accessed through the FluView Interactive website (*19*).

#### Estimating the impact of cross immunity and seasonal drivers of transmission

We used the effective reproduction number, defined as the average number of secondary infections caused by a single infected individual, to quantify the time-varying transmissibility of each coronavirus strain. We estimated the daily effective reproduction number (*R*_*u*_) based on case counts and the generation interval distribution (*20*), with the following parameterization (*21*):

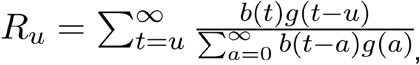

where *b(t)* is the strain-level incidence proxy on day *t*, and *g(a)* is the value of the generation interval distribution at time *a*. To translate the weekly incidence proxies to measures of daily incidence, we followed a previously described spline-based procedure (*22*). For smoothing, the final weekly R is the geometric mean of the daily values in the preceding, current, and following weeks (moving 3-week average). We discarded the first three and last three weekly R estimates. To avoid unstable R estimates resulting from periods of low coronavirus activity, we limited our analysis to “in-season” estimates. Seasons were defined as epidemiological week 40 of each year through week 20 of the following year (roughly October through May). Because 2014 consisted of 53 weeks, we truncated the 2014-15 season at week 19 of 2015.

The generation interval for the four commonly circulating coronaviruses has not been well-studied. In this analysis, we used the estimated serial interval distribution for SARS (Weibull with mean 8.4 days and standard deviation 3.8 days (*32*)) and varied this assumption in sensitivity analyses based on observations that the serial interval for currently circulating human coronaviruses and SARS-CoV-2 may be considerably lower (*33*). We assumed that the maximum generation interval was the first day at which over 99% of the density had been captured.

To understand the relative contribution of depletion of susceptibles compared to seasonal forcing in the observed data, we adapted a linear regression model as follows (*22*):

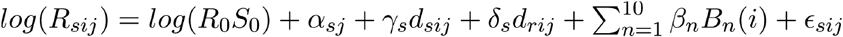

where *R*_*sij*_ is the weekly effective reproduction number for strain *s* in week *i* of season *j, R*_*0*_ is the basic reproduction number, *S*_*0*_ is the fraction of susceptibles at the start of the first season for the reference strain, and ∈_*sij*_ is a normally distributed error term. A dummy variable for each strain-season combination (α_sj_) captures differences in the reproduction number and the starting fraction of susceptibles between strains and over time, but is unable to distinguish between the two. The next two terms estimate the impact of depletion of susceptibles due to infection by the same strain (*d*_*sij*_) and the other betacoronavirus strain (*d*_*rij*_). Depletion of susceptibles for each strain was estimated up to a proportionality constant by the cumulative sum of the incidence proxy over season *j* through week *i*. The coefficient on the first term (γ_s_) represents the scaling factor between the cumulative incidence proxy and true depletion of susceptibles, while δ_s_ captures the level of cross-immunity in addition to scaling; both coefficients were allowed to vary by strain. Because specific seasonal drivers (e.g. absolute humidity) of seasonal variation in coronavirus transmission have not been identified, we did not include them in our model but used a cubic basis spline with knots every four weeks to capture fluctuations in seasonal forcing over the year. *B*_*n*_ represents the ten basis functions for a cubic spline with seven internal knots and no intercept, and β_n_ the coefficients corresponding to each of these functions.

#### Dynamic transmission model

We implemented a two-strain ordinary differential equation (ODE) susceptible-exposed-infectious-recovered (SEIR) compartmental model to describe the transmission dynamics of HCoV-OC43 (‘strain 1’) and HCoV-HKU1 (‘strain 2’) in the United States. Exposed individuals became infectious at rate *ν* and infectious individuals recovered at rate *γ*. Immunity waned at rates *σ*_*1*_ and *σ*_*2*_ for HCoV-OC43 and HCoV-HKU1, respectively. The basic reproduction number, *R*_*0*_, was assumed to be seasonal with a period of 52 weeks, specified by the equation

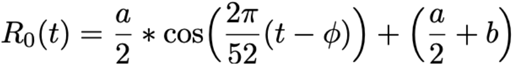

where *a* is the amplitude of the seasonal forcing term, *b* is the minimum value, and *Φ* is the phase shift in weeks. The transmission rate *β(t)* is related to the basic reproduction number by the formula (*11*)

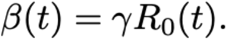

Cross immunity of HCoV-OC43 against HCoV-HKU1 was captured by *χ*_*1,2*_ such that the transmission rate of HCoV-HKU1 to an individual who is exposed to, infected with, or recovered from HCoV-OC43 was reduced by a factor of *1-χ*_*1,2*_, and vice-versa. Individuals died at rate *μ* such that the average lifespan was 1/*μ* = 80 years. Fully-susceptible individuals were born at the same rate *μ* to keep the population size constant. A schematic of the model structure is depicted in **Fig S3**.

The entire population was assumed to be susceptible at the start of the simulation period (time = 0). Infection was introduced through a brief, small pulse in the force of infection (an increase of 0.01/week for one half week) for each strain within the first year of the simulation, simulating the establishment of sustained person-to-person transmission. The model was run for 24.5 years to allow the dynamics to reach a steady state, and then the simulated incidence of Strain 1 and Strain 2 were compared with the percent test-positives multiplied by percent of clinic visits for ILI for HCoV-OC43 and HCoV-HKU1, respectively. Model fit was assessed by log likelihood. To determine the parameter values consistent with HCoV-OC43 and HCoV-HKU1 transmission, we used latin hypercube sampling (LHS) (*34*) to simulate transmission for 100,000 combinations of the model parameters sampled uniformly from the ranges reported in Table 1.

We then used a hill-climbing algorithm to identify the maximum likelihood parameter values, using the best-fit parameter combination from the LHS scheme as initial conditions. The model was implemented in *R* version 3.6.1 (*35*) and solved using the *lsoda()* function (*36*). We further validated the model by visually comparing its estimates of the effective reproduction number for each strain against the regression-based estimates described above. The model-based effective reproduction number was calculated as the product of the basic reproduction number and the proportion of susceptible individuals in the population at time *t*, accounting for cross-immunity (*37*).

Next, we incorporated a third strain into the dynamic transmission model to represent SARS-CoV-2. Using the maximum likelihood parameter values, we simulated transmission of HCoV-OC43 and HCoV-HKU1 for 20 years and then simulated the establishment of sustained SARS-CoV-2 transmission using another half-week pulse in the force of infection. We assumed that the incubation period and infectious period for SARS-CoV-2 were the same as the best-fit values for the other betacoronaviruses (5.0 and 4.9 days, respectively; see Table S7), in broad agreement with other estimates (*23*–*25*). We allowed the cross immunities, duration of immunity, degree of seasonal variation in R0, and establishment time of SARS-CoV-2 to vary. In particular, we allowed the cross immunity from SARS-CoV-2 to the other betacoronaviruses to range from 0 to 1, the cross immunity from the other betacoronaviruses to SARS-CoV-2 to range from 0 to 0.5 (following the observation that SARS infection can induce long-lasting neutralizing antibodies against HCoV-OC43 but not vice-versa (*15*)),the duration of immunity to SARS-CoV-2 to range from 40 weeks to permanent, the seasonal variation in *R*_*0*_ to vary between none and equivalent to the other human betacoronaviruses, and the establishment time to vary throughout 2020. To adjust the amount of seasonal variation in *R*_*0*_, we held the maximum wintertime value of the sinusoid fixed and adjusted the minimum summertime (baseline) value. This way, smaller degrees of seasonal forcing translated into smaller summertime declines in *R*_*0*_; for the no-seasonality scenario, *R*_*0*_ was therefore held fixed at its maximal wintertime value. This choice was informed by observations on the seasonal variation in *R*_*0*_ for influenza, for which the wintertime *R*_*0*_ was similar between geographic locations with distinct climates, while the summertime *R*_*0*_ varied substantially between locations (*11*). For a representative set of parameter values within these ranges, we measured the annual incidence of infection due to SARS-CoV-2 and the annual SARS-CoV-2 outbreak peak size for the five years following the simulated time of establishment. We summarized the post-pandemic SARS-CoV-2 dynamics into the categories of annual outbreaks, biennial outbreaks, sporadic outbreaks, or virtual elimination.

---

**Table S1.** Estimated regression model coefficients.

| Parameter | Strain | Season | Value |
| --- | --- | --- | --- |
| Intercept | HCoV-HKU1 | 2014-15 | 0.1277 |
| $\alpha_{sj}$ | HCoV-HKU1 | 2015-16 | -0.1114 |
| $\alpha_{sj}$ | HCoV-HKU1 | 2016-17 | 0.0223 |
| $\alpha_{sj}$ | HCoV-HKU1 | 2017-18 | 0.0799 |
| $\alpha_{sj}$ | HCoV-HKU1 | 2018-19 | 0.0651 |
| $\alpha_{sj}$ | HCoV-OC43 | 2014-15 | -0.0627 |
| $\alpha_{sj}$ | HCoV-OC43 | 2015-16 | 0.0416 |
| $\alpha_{sj}$ | HCoV-OC43 | 2016-17 | -0.0620 |
| $\alpha_{sj}$ | HCoV-OC43 | 2017-18 | -0.0716 |
| $\alpha_{sj}$ | HCoV-OC43 | 2018-19 | -0.0566 |
| $\gamma_s$ (reference) | HCoV-HKU1 | - | -0.0024 |
| $\gamma_s$ | HCoV-OC43 | - | 0.0009 |
| $\delta_s$ (reference) | HCoV-HKU1 | - | -0.0012 |
| $\delta_s$ | HCoV-OC43 | - | 0.0005 |
| $\beta_1$ | - | - | 0.4513 |
| $\beta_2$ | - | - | 0.2444 |
| $\beta_3$ | - | - | 0.4803 |
| $\beta_4$ | - | - | 0.0644 |
| $\beta_5$ | - | - | 0.3300 |
| $\beta_6$ | - | - | 0.0057 |
| $\beta_7$ | - | - | 0.1105 |
| $\beta_8$ | - | - | 0.0539 |
| $\beta_9$ | - | - | 0.2128 |
| $\beta_{10}$ | - | - | 0.2522 |

**Table S2.** Cumulative projected percent influenza-like illness (ILI) x percent positive laboratory tests for SARS-CoV-2 by year for a representative set of cross immunities, immunity durations, and establishment times. Percent ILI is measured as the population-weighted proportion of visits to sentinel providers that were associated with influenza symptoms. The seasonality factor represents the amount of seasonal variation in the SARS-CoV-2 *R*_*0*_ relative to the other human betacoronaviruses. A seasonality factor of 0.5 indicates that the amplitude of seasonal variation in *R*_*0*_ for SARS-CoV-2 is half the amplitude of the seasonal variation in *R*_*0*_ for the other betacoronaviruses. Chi3X represents the degree of cross-immunity induced by infection with SARS-CoV-2 against OC43 and HKU1 and ChiX3 represents the degree of cross-immunity induced by OC43 or HKU1 infection against SARS-CoV-2. The establishment times correspond to: Winter - week 4 (early February); Spring - week 16 (late April); Summer - week 28 (mid July); Autumn-week 40 (early October). Darker shading corresponds to higher cumulative infection sizes.

| Chi3X | ChiX3 | Immunity (wks) | Est. time | Seasonality factor | 2019 | 2020 | 2021 | 2022 | 2023 | 2024 | 2025 | Max | Post-pandemic transmission |
| --- | --- | --- | --- | --- | --- | --- | --- | --- | --- | --- | --- | --- | --- |
| 0.7 | 0 | 40 | Winter | 0.5 | 0.00 | 6.05 | 3.54 | 4.21 | 4.43 | 4.47 | 4.47 | 27.17 | Annual |
| 0.7 | 0.3 | 40 | Winter | 0.5 | 0.00 | 4.18 | 3.94 | 4.38 | 4.22 | 4.00 | 4.26 | 24.97 | Annual |
| 0.3 | 0 | 40 | Winter | 0.5 | 0.00 | 6.05 | 3.54 | 4.21 | 4.43 | 4.47 | 4.47 | 27.17 | Annual |
| 0 | 0 | 40 | Winter | 0.5 | 0.00 | 6.05 | 3.54 | 4.21 | 4.43 | 4.47 | 4.47 | 27.17 | Annual |
| 0.7 | 0 | 104 | Winter | 0.5 | 0.00 | 5.73 | 0.04 | 1.93 | 1.73 | 2.14 | 1.43 | 13.01 | Annual |
| 0.7 | 0.3 | 104 | Winter | 0.5 | 0.00 | 3.96 | 0.96 | 2.76 | 0.69 | 2.27 | 1.20 | 11.83 | Annual |
| 0.3 | 0 | 104 | Winter | 0.5 | 0.00 | 5.73 | 0.04 | 1.93 | 1.74 | 2.14 | 1.43 | 13.01 | Annual |
| 0 | 0 | 104 | Winter | 0.5 | 0.00 | 5.73 | 0.04 | 1.93 | 1.74 | 2.14 | 1.43 | 13.01 | Annual |
| 0.7 | 0 | Permanent | Winter | 0.5 | 0.00 | 5.55 | 0.01 | 0.00 | 0.00 | 0.00 | 0.00 | 5.56 | Virtual elimination |
| 0.7 | 0.3 | Permanent | Winter | 0.5 | 0.00 | 3.82 | 0.33 | 0.00 | 0.00 | 0.00 | 0.00 | 4.15 | Virtual elimination |
| 0.3 | 0 | Permanent | Winter | 0.5 | 0.00 | 5.55 | 0.01 | 0.00 | 0.00 | 0.00 | 0.00 | 5.56 | Virtual elimination |
| 0 | 0 | Permanent | Winter | 0.5 | 0.00 | 5.55 | 0.01 | 0.00 | 0.00 | 0.00 | 0.00 | 5.56 | Virtual elimination |
| 0.7 | 0 | 40 | Spring | 0.5 | 0.00 | 2.54 | 5.28 | 4.67 | 4.56 | 4.52 | 4.49 | 26.05 | Annual |
| 0.7 | 0.3 | 40 | Spring | 0.5 | 0.00 | 0.88 | 5.86 | 4.73 | 4.23 | 4.16 | 4.14 | 24.00 | Annual |
| 0.3 | 0 | 40 | Spring | 0.5 | 0.00 | 2.54 | 5.28 | 4.67 | 4.56 | 4.52 | 4.49 | 26.05 | Annual |
| 0 | 0 | 40 | Spring | 0.5 | 0.00 | 2.54 | 5.28 | 4.67 | 4.56 | 4.52 | 4.49 | 26.05 | Annual |
| 0.7 | 0 | 104 | Spring | 0.5 | 0.00 | 2.52 | 3.22 | 0.65 | 2.93 | 1.34 | 2.12 | 12.77 | Annual |
| 0.7 | 0.3 | 104 | Spring | 0.5 | 0.00 | 0.88 | 4.31 | 1.49 | 1.87 | 1.28 | 2.09 | 11.92 | Annual |
| 0.3 | 0 | 104 | Spring | 0.5 | 0.00 | 2.52 | 3.22 | 0.65 | 2.93 | 1.34 | 2.12 | 12.77 | Annual |
| 0 | 0 | 104 | Spring | 0.5 | 0.00 | 2.52 | 3.22 | 0.65 | 2.93 | 1.34 | 2.12 | 12.77 | Annual |
| 0.7 | 0 | Permanent | Spring | 0.5 | 0.00 | 2.51 | 2.94 | 0.00 | 0.00 | 0.00 | 0.00 | 5.45 | Virtual elimination |
| 0.7 | 0.3 | Permanent | Spring | 0.5 | 0.00 | 0.87 | 3.86 | 0.00 | 0.00 | 0.00 | 0.00 | 4.73 | Virtual elimination |
| 0.3 | 0 | Permanent | Spring | 0.5 | 0.00 | 2.51 | 2.94 | 0.00 | 0.00 | 0.00 | 0.00 | 5.45 | Virtual elimination |
| 0 | 0 | Permanent | Spring | 0.5 | 0.00 | 2.51 | 2.94 | 0.00 | 0.00 | 0.00 | 0.00 | 5.45 | Virtual elimination |
| 0.7 | 0 | 40 | Summer | 0.5 | 0.00 | 0.00 | 6.76 | 4.84 | 4.54 | 4.46 | 4.45 | 25.05 | Annual |
| 0.7 | 0.3 | 40 | Summer | 0.5 | 0.00 | 0.00 | 6.27 | 4.69 | 3.89 | 4.34 | 3.90 | 23.09 | Annual |
| 0.3 | 0 | 40 | Summer | 0.5 | 0.00 | 0.00 | 6.76 | 4.84 | 4.54 | 4.46 | 4.45 | 25.05 | Annual |
| 0 | 0 | 40 | Summer | 0.5 | 0.00 | 0.00 | 6.76 | 4.84 | 4.54 | 4.46 | 4.45 | 25.05 | Annual |
| 0.7 | 0 | 104 | Summer | 0.5 | 0.00 | 0.00 | 6.14 | 0.00 | 3.30 | 0.34 | 2.92 | 12.71 | Biennial |
| 0.7 | 0.3 | 104 | Summer | 0.5 | 0.00 | 0.00 | 5.61 | 0.02 | 2.74 | 0.80 | 2.54 | 11.70 | Biennial |
| 0.3 | 0 | 104 | Summer | 0.5 | 0.00 | 0.00 | 6.14 | 0.00 | 3.30 | 0.34 | 2.92 | 12.71 | Biennial |
| 0 | 0 | 104 | Summer | 0.5 | 0.00 | 0.00 | 6.14 | 0.00 | 3.30 | 0.34 | 2.92 | 12.71 | Biennial |
| 0.7 | 0 | Permanent | Summer | 0.5 | 0.00 | 0.00 | 5.90 | 0.00 | 0.00 | 0.00 | 0.00 | 5.90 | Virtual elimination |
| 0.7 | 0.3 | Permanent | Summer | 0.5 | 0.00 | 0.00 | 5.32 | 0.00 | 0.00 | 0.00 | 0.00 | 5.32 | Virtual elimination |
| 0.3 | 0 | Permanent | Summer | 0.5 | 0.00 | 0.00 | 5.90 | 0.00 | 0.00 | 0.00 | 0.00 | 5.90 | Virtual elimination |
| 0 | 0 | Permanent | Summer | 0.5 | 0.00 | 0.00 | 5.90 | 0.00 | 0.00 | 0.00 | 0.00 | 5.90 | Virtual elimination |
| 0.7 | 0 | 40 | Autumn | 0.5 | 0.00 | 0.00 | 6.68 | 4.41 | 4.23 | 4.37 | 4.43 | 24.11 | Annual |
| 0.7 | 0.3 | 40 | Autumn | 0.5 | 0.00 | 0.00 | 6.09 | 4.21 | 3.73 | 4.21 | 3.97 | 22.20 | Annual |
| 0.3 | 0 | 40 | Autumn | 0.5 | 0.00 | 0.00 | 6.68 | 4.41 | 4.23 | 4.37 | 4.43 | 24.11 | Annual |
| 0 | 0 | 40 | Autumn | 0.5 | 0.00 | 0.00 | 6.68 | 4.41 | 4.23 | 4.37 | 4.43 | 24.11 | Annual |
| 0.7 | 0 | 104 | Autumn | 0.5 | 0.00 | 0.00 | 6.29 | 0.00 | 0.68 | 3.25 | 1.38 | 11.59 | Sporadic |
| 0.7 | 0.3 | 104 | Autumn | 0.5 | 0.00 | 0.00 | 5.68 | 0.00 | 1.41 | 2.46 | 1.54 | 11.09 | Sporadic |
| 0.3 | 0 | 104 | Autumn | 0.5 | 0.00 | 0.00 | 6.29 | 0.00 | 0.67 | 3.25 | 1.37 | 11.59 | Sporadic |
| 0 | 0 | 104 | Autumn | 0.5 | 0.00 | 0.00 | 6.29 | 0.00 | 0.67 | 3.25 | 1.37 | 11.59 | Sporadic |
| 0.7 | 0 | Permanent | Autumn | 0.5 | 0.00 | 0.00 | 6.09 | 0.00 | 0.00 | 0.00 | 0.00 | 6.09 | Virtual elimination |
| 0.7 | 0.3 | Permanent | Autumn | 0.5 | 0.00 | 0.00 | 5.47 | 0.00 | 0.00 | 0.00 | 0.00 | 5.47 | Virtual elimination |
| 0.3 | 0 | Permanent | Autumn | 0.5 | 0.00 | 0.00 | 6.09 | 0.00 | 0.00 | 0.00 | 0.00 | 6.09 | Virtual elimination |
| 0 | 0 | Permanent | Autumn | 0.5 | 0.00 | 0.00 | 6.09 | 0.00 | 0.00 | 0.00 | 0.00 | 6.09 | Virtual elimination |

**Table S3.** Cumulative projected percent influenza-like illness (ILI) x percent positive laboratory tests for SARS-CoV-2 by year for a representative set of cross immunities, immunity durations, and establishment times. Percent ILI is measured as the population-weighted proportion of visits to sentinel providers that were associated with influenza symptoms. The seasonality factor represents the amount of seasonal variation in the SARS-CoV-2 *R*_*0*_ relative to the other human betacoronaviruses. A seasonality factor of 0 indicates no seasonal variation in *R*_*0*_ for SARS-CoV-2. Chi3X represents the degree of cross-immunity induced by infection with SARS-CoV-2 against OC43 and HKU1 and ChiX3 represents the degree of cross-immunity induced by OC43 or HKU1 infection against SARS-CoV-2. The establishment times correspond to: Winter - week 4 (early February); Spring - week 16 (late April); Summer - week 28 (mid July); Autumn - week 40 (early October). Darker shading corresponds to higher cumulative infection sizes.

| Chi3X | ChiX3 | Immunity (wks) | Est. time | Seasonality factor | 2019 | 2020 | 2021 | 2022 | 2023 | 2024 | 2025 | Max | Post-pandemic transmission |
| --- | --- | --- | --- | --- | --- | --- | --- | --- | --- | --- | --- | --- | --- |
| 0.7 | 0 | 40 | Winter | 0 | 0.00 | 6.65 | 4.11 | 4.50 | 4.76 | 4.87 | 4.89 | 29.79 | Annual |
| 0.7 | 0.3 | 40 | Winter | 0 | 0.00 | 5.44 | 4.11 | 4.61 | 4.77 | 4.62 | 4.50 | 28.05 | Annual |
| 0.3 | 0 | 40 | Winter | 0 | 0.00 | 6.65 | 4.11 | 4.50 | 4.76 | 4.87 | 4.89 | 29.79 | Annual |
| 0 | 0 | 40 | Winter | 0 | 0.00 | 6.65 | 4.11 | 4.50 | 4.76 | 4.87 | 4.89 | 29.79 | Annual |
| 0.7 | 0 | 104 | Winter | 0 | 0.00 | 6.30 | 0.01 | 2.11 | 1.50 | 2.67 | 1.48 | 14.08 | Annual |
| 0.7 | 0.3 | 104 | Winter | 0 | 0.00 | 5.12 | 0.23 | 3.08 | 1.24 | 1.92 | 1.88 | 13.48 | Annual |
| 0.3 | 0 | 104 | Winter | 0 | 0.00 | 6.30 | 0.01 | 2.10 | 1.51 | 2.67 | 1.48 | 14.08 | Annual |
| 0 | 0 | 104 | Winter | 0 | 0.00 | 6.30 | 0.01 | 2.10 | 1.51 | 2.67 | 1.48 | 14.08 | Annual |
| 0.7 | 0 | Permanent | Winter | 0 | 0.00 | 6.11 | 0.00 | 0.00 | 0.00 | 0.00 | 0.00 | 6.11 | Virtual elimination |
| 0.7 | 0.3 | Permanent | Winter | 0 | 0.00 | 4.93 | 0.11 | 0.00 | 0.00 | 0.00 | 0.00 | 5.04 | Virtual elimination |
| 0.3 | 0 | Permanent | Winter | 0 | 0.00 | 6.11 | 0.00 | 0.00 | 0.00 | 0.00 | 0.00 | 6.11 | Virtual elimination |
| 0 | 0 | Permanent | Winter | 0 | 0.00 | 6.11 | 0.00 | 0.00 | 0.00 | 0.00 | 0.00 | 6.11 | Virtual elimination |
| 0.7 | 0 | 40 | Spring | 0 | 0.00 | 4.50 | 4.47 | 5.15 | 4.83 | 4.84 | 4.87 | 28.67 | Annual |
| 0.7 | 0.3 | 40 | Spring | 0 | 0.00 | 1.97 | 5.93 | 5.25 | 4.87 | 4.52 | 4.59 | 27.13 | Annual |
| 0.3 | 0 | 40 | Spring | 0 | 0.00 | 4.50 | 4.47 | 5.15 | 4.83 | 4.84 | 4.87 | 28.67 | Annual |
| 0 | 0 | 40 | Spring | 0 | 0.00 | 4.50 | 4.47 | 5.15 | 4.83 | 4.84 | 4.87 | 28.67 | Annual |
| 0.7 | 0 | 104 | Spring | 0 | 0.00 | 4.44 | 1.88 | 0.20 | 3.36 | 2.31 | 1.20 | 13.39 | Annual |
| 0.7 | 0.3 | 104 | Spring | 0 | 0.00 | 1.96 | 3.58 | 2.55 | 0.95 | 2.40 | 1.70 | 13.13 | Annual |
| 0.3 | 0 | 104 | Spring | 0 | 0.00 | 4.44 | 1.88 | 0.20 | 3.36 | 2.31 | 1.20 | 13.38 | Annual |
| 0 | 0 | 104 | Spring | 0 | 0.00 | 4.44 | 1.88 | 0.20 | 3.37 | 2.30 | 1.20 | 13.38 | Annual |
| 0.7 | 0 | Permanent | Spring | 0 | 0.00 | 4.40 | 1.70 | 0.00 | 0.00 | 0.00 | 0.00 | 6.11 | Virtual elimination |
| 0.7 | 0.3 | Permanent | Spring | 0 | 0.00 | 1.95 | 3.32 | 0.00 | 0.00 | 0.00 | 0.00 | 5.27 | Virtual elimination |
| 0.3 | 0 | Permanent | Spring | 0 | 0.00 | 4.40 | 1.70 | 0.00 | 0.00 | 0.00 | 0.00 | 6.11 | Virtual elimination |
| 0 | 0 | Permanent | Spring | 0 | 0.00 | 4.40 | 1.70 | 0.00 | 0.00 | 0.00 | 0.00 | 6.11 | Virtual elimination |
| 0.7 | 0 | 40 | Summer | 0 | 0.00 | 0.00 | 7.05 | 5.52 | 5.15 | 4.94 | 4.89 | 27.54 | Annual |
| 0.7 | 0.3 | 40 | Summer | 0 | 0.00 | 0.00 | 6.56 | 5.36 | 4.99 | 4.49 | 4.75 | 26.15 | Annual |
| 0.3 | 0 | 40 | Summer | 0 | 0.00 | 0.00 | 7.05 | 5.52 | 5.15 | 4.94 | 4.89 | 27.54 | Annual |
| 0 | 0 | 40 | Summer | 0 | 0.00 | 0.00 | 7.05 | 5.52 | 5.15 | 4.94 | 4.89 | 27.54 | Annual |
| 0.7 | 0 | 104 | Summer | 0 | 0.00 | 0.00 | 6.32 | 0.01 | 3.51 | 1.29 | 1.89 | 13.02 | Annual |
| 0.7 | 0.3 | 104 | Summer | 0 | 0.00 | 0.00 | 5.72 | 0.36 | 3.00 | 1.70 | 1.95 | 12.74 | Annual |
| 0.3 | 0 | 104 | Summer | 0 | 0.00 | 0.00 | 6.32 | 0.01 | 3.51 | 1.28 | 1.91 | 13.02 | Annual |
| 0 | 0 | 104 | Summer | 0 | 0.00 | 0.00 | 6.32 | 0.01 | 3.51 | 1.28 | 1.91 | 13.02 | Annual |
| 0.7 | 0 | Permanent | Summer | 0 | 0.00 | 0.00 | 6.11 | 0.00 | 0.00 | 0.00 | 0.00 | 6.11 | Virtual elimination |
| 0.7 | 0.3 | Permanent | Summer | 0 | 0.00 | 0.00 | 5.47 | 0.00 | 0.00 | 0.00 | 0.00 | 5.47 | Virtual elimination |
| 0.3 | 0 | Permanent | Summer | 0 | 0.00 | 0.00 | 6.11 | 0.00 | 0.00 | 0.00 | 0.00 | 6.11 | Virtual elimination |
| 0 | 0 | Permanent | Summer | 0 | 0.00 | 0.00 | 6.11 | 0.00 | 0.00 | 0.00 | 0.00 | 6.11 | Virtual elimination |
| 0.7 | 0 | 40 | Autumn | 0 | 0.00 | 0.00 | 6.78 | 4.80 | 4.93 | 4.97 | 4.92 | 26.40 | Annual |
| 0.7 | 0.3 | 40 | Autumn | 0 | 0.00 | 0.00 | 6.26 | 4.73 | 4.73 | 4.50 | 4.74 | 24.96 | Annual |
| 0.3 | 0 | 40 | Autumn | 0 | 0.00 | 0.00 | 6.78 | 4.80 | 4.93 | 4.97 | 4.92 | 26.40 | Annual |
| 0 | 0 | 40 | Autumn | 0 | 0.00 | 0.00 | 6.78 | 4.80 | 4.93 | 4.97 | 4.92 | 26.40 | Annual |
| 0.7 | 0 | 104 | Autumn | 0 | 0.00 | 0.00 | 6.32 | 0.00 | 3.43 | 0.46 | 2.63 | 12.84 | Biennial |
| 0.7 | 0.3 | 104 | Autumn | 0 | 0.00 | 0.00 | 5.75 | 0.03 | 3.24 | 1.00 | 2.12 | 12.14 | Biennial |
| 0.3 | 0 | 104 | Autumn | 0 | 0.00 | 0.00 | 6.32 | 0.00 | 3.43 | 0.46 | 2.63 | 12.84 | Biennial |
| 0 | 0 | 104 | Autumn | 0 | 0.00 | 0.00 | 6.32 | 0.00 | 3.43 | 0.46 | 2.63 | 12.84 | Biennial |
| 0.7 | 0 | Permanent | Autumn | 0 | 0.00 | 0.00 | 6.11 | 0.00 | 0.00 | 0.00 | 0.00 | 6.11 | Virtual elimination |
| 0.7 | 0.3 | Permanent | Autumn | 0 | 0.00 | 0.00 | 5.52 | 0.00 | 0.00 | 0.00 | 0.00 | 5.52 | Virtual elimination |
| 0.3 | 0 | Permanent | Autumn | 0 | 0.00 | 0.00 | 6.11 | 0.00 | 0.00 | 0.00 | 0.00 | 6.11 | Virtual elimination |
| 0 | 0 | Permanent | Autumn | 0 | 0.00 | 0.00 | 6.11 | 0.00 | 0.00 | 0.00 | 0.00 | 6.11 | Virtual elimination |

**Table S4.**
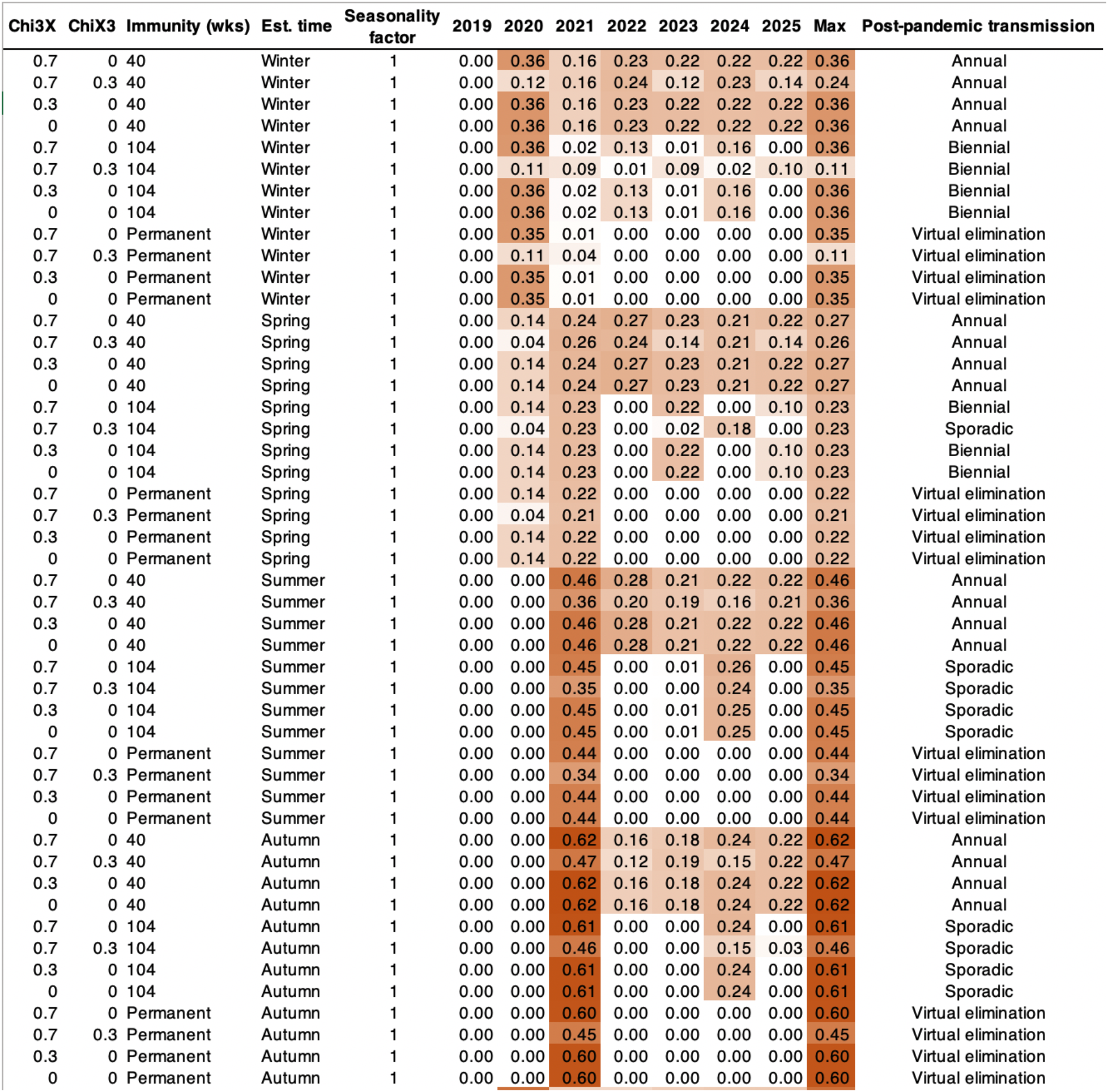
Peak simulated SARS-CoV-2 epidemic sizes, in units of percent influenza-like illness (ILI) x percent positive laboratory tests, by year for a representative set of cross immunities, immunity durations, and establishment times. Percent ILI is measured as the population-weighted proportion of visits to sentinel providers that were associated with influenza symptoms. The seasonality factor represents the amount of seasonal variation in the SARS-CoV-2 *R*_*0*_ relative to the other human betacoronaviruses. A seasonality factor of 1 indicates equal seasonal variation in *R*_*0*_ for SARS-CoV-2 as for the other human betacoronaviruses. Chi3X represents the degree of cross-immunity induced by infection with SARS-CoV-2 against OC43 and HKU1 and ChiX3 represents the degree of cross-immunity induced by OC43 or HKU1 infection against SARS-CoV-2. The establishment times correspond to: Winter - week 4 (early February); Spring - week 16 (late April); Summer - week 28 (mid July); Autumn - week 40 (early October). Darker shading corresponds to higher peak sizes.

**Table S5.** Peak simulated SARS-CoV-2 epidemic sizes, in units of percent influenza-like illness (ILI) x percent positive laboratory tests, by year for a representative set of cross immunities, immunity durations, and establishment times. Percent ILI is measured as the population-weighted proportion of visits to sentinel providers that were associated with influenza symptoms. The seasonality factor represents the amount of seasonal variation in the SARS-CoV-2 *R*_*0*_ relative to the other human betacoronaviruses. A seasonality factor of 0.5 indicates that the amplitude of seasonal variation in *R*_*0*_ for SARS-CoV-2 is half the amplitude of the seasonal variation in *R*_*0*_ for the other betacoronaviruses. Chi3X represents the degree of cross-immunity induced by infection with SARS-CoV-2 against OC43 and HKU1 and ChiX3 represents the degree of cross-immunity induced by OC43 or HKU1 infection against SARS-CoV-2. The establishment times correspond to: Winter - week 4 (early February); Spring - week 16 (late April); Summer - week 28 (mid July); Autumn - week 40 (early October). Darker shading corresponds to higher peak sizes.

| Chi3X | ChiX3 | Immunity (wks) | Est. time | Seasonality factor | 2019 | 2020 | 2021 | 2022 | 2023 | 2024 | 2025 | Max | Post-pandemic transmission |
| --- | --- | --- | --- | --- | --- | --- | --- | --- | --- | --- | --- | --- | --- |
| 0.7 | 0 | 40 | Winter | 0.5 | 0.00 | 0.50 | 0.14 | 0.13 | 0.16 | 0.18 | 0.18 | 0.50 | Annual |
| 0.7 | 0.3 | 40 | Winter | 0.5 | 0.00 | 0.23 | 0.10 | 0.14 | 0.17 | 0.17 | 0.18 | 0.23 | Annual |
| 0.3 | 0 | 40 | Winter | 0.5 | 0.00 | 0.50 | 0.14 | 0.13 | 0.16 | 0.18 | 0.18 | 0.50 | Annual |
| 0 | 0 | 40 | Winter | 0.5 | 0.00 | 0.50 | 0.14 | 0.13 | 0.16 | 0.18 | 0.18 | 0.50 | Annual |
| 0.7 | 0 | 104 | Winter | 0.5 | 0.00 | 0.49 | 0.01 | 0.09 | 0.06 | 0.07 | 0.04 | 0.49 | Annual |
| 0.7 | 0.3 | 104 | Winter | 0.5 | 0.00 | 0.22 | 0.07 | 0.09 | 0.02 | 0.07 | 0.04 | 0.22 | Annual |
| 0.3 | 0 | 104 | Winter | 0.5 | 0.00 | 0.49 | 0.01 | 0.09 | 0.06 | 0.07 | 0.04 | 0.49 | Annual |
| 0 | 0 | 104 | Winter | 0.5 | 0.00 | 0.49 | 0.01 | 0.09 | 0.06 | 0.07 | 0.04 | 0.49 | Annual |
| 0.7 | 0 | Permanent | Winter | 0.5 | 0.00 | 0.49 | 0.00 | 0.00 | 0.00 | 0.00 | 0.00 | 0.49 | Virtual elimination |
| 0.7 | 0.3 | Permanent | Winter | 0.5 | 0.00 | 0.22 | 0.06 | 0.00 | 0.00 | 0.00 | 0.00 | 0.22 | Virtual elimination |
| 0.3 | 0 | Permanent | Winter | 0.5 | 0.00 | 0.49 | 0.00 | 0.00 | 0.00 | 0.00 | 0.00 | 0.49 | Virtual elimination |
| 0 | 0 | Permanent | Winter | 0.5 | 0.00 | 0.49 | 0.00 | 0.00 | 0.00 | 0.00 | 0.00 | 0.49 | Virtual elimination |
| 0.7 | 0 | 40 | Spring | 0.5 | 0.00 | 0.40 | 0.42 | 0.11 | 0.16 | 0.18 | 0.18 | 0.42 | Annual |
| 0.7 | 0.3 | 40 | Spring | 0.5 | 0.00 | 0.13 | 0.25 | 0.15 | 0.17 | 0.18 | 0.18 | 0.25 | Annual |
| 0.3 | 0 | 40 | Spring | 0.5 | 0.00 | 0.40 | 0.42 | 0.11 | 0.16 | 0.18 | 0.18 | 0.42 | Annual |
| 0 | 0 | 40 | Spring | 0.5 | 0.00 | 0.40 | 0.42 | 0.11 | 0.16 | 0.18 | 0.18 | 0.42 | Annual |
| 0.7 | 0 | 104 | Spring | 0.5 | 0.00 | 0.40 | 0.41 | 0.05 | 0.08 | 0.05 | 0.04 | 0.41 | Annual |
| 0.7 | 0.3 | 104 | Spring | 0.5 | 0.00 | 0.13 | 0.24 | 0.06 | 0.05 | 0.04 | 0.06 | 0.24 | Annual |
| 0.3 | 0 | 104 | Spring | 0.5 | 0.00 | 0.40 | 0.41 | 0.05 | 0.08 | 0.05 | 0.04 | 0.41 | Annual |
| 0 | 0 | 104 | Spring | 0.5 | 0.00 | 0.40 | 0.41 | 0.05 | 0.08 | 0.05 | 0.04 | 0.41 | Annual |
| 0.7 | 0 | Permanent | Spring | 0.5 | 0.00 | 0.39 | 0.40 | 0.00 | 0.00 | 0.00 | 0.00 | 0.40 | Virtual elimination |
| 0.7 | 0.3 | Permanent | Spring | 0.5 | 0.00 | 0.13 | 0.23 | 0.00 | 0.00 | 0.00 | 0.00 | 0.23 | Virtual elimination |
| 0.3 | 0 | Permanent | Spring | 0.5 | 0.00 | 0.39 | 0.40 | 0.00 | 0.00 | 0.00 | 0.00 | 0.40 | Virtual elimination |
| 0 | 0 | Permanent | Spring | 0.5 | 0.00 | 0.39 | 0.40 | 0.00 | 0.00 | 0.00 | 0.00 | 0.40 | Virtual elimination |
| 0.7 | 0 | 40 | Summer | 0.5 | 0.00 | 0.00 | 0.53 | 0.22 | 0.19 | 0.19 | 0.18 | 0.53 | Annual |
| 0.7 | 0.3 | 40 | Summer | 0.5 | 0.00 | 0.00 | 0.39 | 0.22 | 0.17 | 0.19 | 0.16 | 0.39 | Annual |
| 0.3 | 0 | 40 | Summer | 0.5 | 0.00 | 0.00 | 0.53 | 0.22 | 0.19 | 0.19 | 0.18 | 0.53 | Annual |
| 0 | 0 | 40 | Summer | 0.5 | 0.00 | 0.00 | 0.53 | 0.22 | 0.19 | 0.19 | 0.18 | 0.53 | Annual |
| 0.7 | 0 | 104 | Summer | 0.5 | 0.00 | 0.00 | 0.52 | 0.00 | 0.17 | 0.01 | 0.10 | 0.52 | Biennial |
| 0.7 | 0.3 | 104 | Summer | 0.5 | 0.00 | 0.00 | 0.37 | 0.00 | 0.13 | 0.04 | 0.06 | 0.37 | Biennial |
| 0.3 | 0 | 104 | Summer | 0.5 | 0.00 | 0.00 | 0.52 | 0.00 | 0.17 | 0.01 | 0.10 | 0.52 | Biennial |
| 0 | 0 | 104 | Summer | 0.5 | 0.00 | 0.00 | 0.52 | 0.00 | 0.17 | 0.01 | 0.10 | 0.52 | Biennial |
| 0.7 | 0 | Permanent | Summer | 0.5 | 0.00 | 0.00 | 0.51 | 0.00 | 0.00 | 0.00 | 0.00 | 0.51 | Virtual elimination |
| 0.7 | 0.3 | Permanent | Summer | 0.5 | 0.00 | 0.00 | 0.37 | 0.00 | 0.00 | 0.00 | 0.00 | 0.37 | Virtual elimination |
| 0.3 | 0 | Permanent | Summer | 0.5 | 0.00 | 0.00 | 0.51 | 0.00 | 0.00 | 0.00 | 0.00 | 0.51 | Virtual elimination |
| 0 | 0 | Permanent | Summer | 0.5 | 0.00 | 0.00 | 0.51 | 0.00 | 0.00 | 0.00 | 0.00 | 0.51 | Virtual elimination |
| 0.7 | 0 | 40 | Autumn | 0.5 | 0.00 | 0.00 | 0.62 | 0.23 | 0.17 | 0.17 | 0.18 | 0.62 | Annual |
| 0.7 | 0.3 | 40 | Autumn | 0.5 | 0.00 | 0.00 | 0.47 | 0.21 | 0.15 | 0.17 | 0.17 | 0.47 | Annual |
| 0.3 | 0 | 40 | Autumn | 0.5 | 0.00 | 0.00 | 0.62 | 0.23 | 0.17 | 0.17 | 0.18 | 0.62 | Annual |
| 0 | 0 | 40 | Autumn | 0.5 | 0.00 | 0.00 | 0.62 | 0.23 | 0.17 | 0.17 | 0.18 | 0.62 | Annual |
| 0.7 | 0 | 104 | Autumn | 0.5 | 0.00 | 0.00 | 0.61 | 0.00 | 0.07 | 0.13 | 0.05 | 0.61 | Sporadic |
| 0.7 | 0.3 | 104 | Autumn | 0.5 | 0.00 | 0.00 | 0.46 | 0.00 | 0.05 | 0.06 | 0.05 | 0.46 | Sporadic |
| 0.3 | 0 | 104 | Autumn | 0.5 | 0.00 | 0.00 | 0.61 | 0.00 | 0.07 | 0.13 | 0.05 | 0.61 | Sporadic |
| 0 | 0 | 104 | Autumn | 0.5 | 0.00 | 0.00 | 0.61 | 0.00 | 0.07 | 0.13 | 0.05 | 0.61 | Sporadic |
| 0.7 | 0 | Permanent | Autumn | 0.5 | 0.00 | 0.00 | 0.61 | 0.00 | 0.00 | 0.00 | 0.00 | 0.61 | Virtual elimination |
| 0.7 | 0.3 | Permanent | Autumn | 0.5 | 0.00 | 0.00 | 0.45 | 0.00 | 0.00 | 0.00 | 0.00 | 0.45 | Virtual elimination |
| 0.3 | 0 | Permanent | Autumn | 0.5 | 0.00 | 0.00 | 0.61 | 0.00 | 0.00 | 0.00 | 0.00 | 0.61 | Virtual elimination |
| 0 | 0 | Permanent | Autumn | 0.5 | 0.00 | 0.00 | 0.61 | 0.00 | 0.00 | 0.00 | 0.00 | 0.61 | Virtual elimination |

**Table S6.** Peak simulated SARS-CoV-2 epidemic sizes, in units of percent influenza-like illness (ILI) x percent positive laboratory tests, by year for a representative set of cross immunities, immunity durations, and establishment times. Percent ILI is measured as the population-weighted proportion of visits to sentinel providers that were associated with influenza symptoms. The seasonality factor represents the amount of seasonal variation in the SARS-CoV-2 *R*_*0*_ relative to the other human betacoronaviruses. A seasonality factor of 0 indicates no seasonal variation in *R*_*0*_ for SARS-CoV-2. Chi3X represents the degree of cross-immunity induced by infection with SARS-CoV-2 against OC43 and HKU1 and ChiX3 represents the degree of cross-immunity induced by OC43 or HKU1 infection against SARS-CoV-2. The establishment times correspond to: Winter - week 4 (early February); Spring - week 16 (late April); Summer - week 28 (mid July); Autumn - week 40 (early October). Darker shading corresponds to higher peak sizes.

| Chi3X | ChiX3 | Immunity (wks) | Est. time | Seasonality factor | 2019 | 2020 | 2021 | 2022 | 2023 | 2024 | 2025 | Max | Post-pandemic transmission |
| --- | --- | --- | --- | --- | --- | --- | --- | --- | --- | --- | --- | --- | --- |
| 0.7 | 0 | 40 | Winter | 0 | 0.00 | 0.63 | 0.17 | 0.09 | 0.08 | 0.07 | 0.07 | 0.63 | Annual |
| 0.7 | 0.3 | 40 | Winter | 0 | 0.00 | 0.35 | 0.13 | 0.09 | 0.07 | 0.07 | 0.07 | 0.35 | Annual |
| 0.3 | 0 | 40 | Winter | 0 | 0.00 | 0.63 | 0.17 | 0.09 | 0.08 | 0.07 | 0.07 | 0.63 | Annual |
| 0 | 0 | 40 | Winter | 0 | 0.00 | 0.63 | 0.17 | 0.09 | 0.08 | 0.07 | 0.07 | 0.63 | Annual |
| 0.7 | 0 | 104 | Winter | 0 | 0.00 | 0.62 | 0.00 | 0.16 | 0.15 | 0.08 | 0.05 | 0.62 | Annual |
| 0.7 | 0.3 | 104 | Winter | 0 | 0.00 | 0.34 | 0.04 | 0.11 | 0.06 | 0.06 | 0.04 | 0.34 | Annual |
| 0.3 | 0 | 104 | Winter | 0 | 0.00 | 0.62 | 0.00 | 0.16 | 0.15 | 0.08 | 0.05 | 0.62 | Annual |
| 0 | 0 | 104 | Winter | 0 | 0.00 | 0.62 | 0.00 | 0.16 | 0.15 | 0.08 | 0.05 | 0.62 | Annual |
| 0.7 | 0 | Permanent | Winter | 0 | 0.00 | 0.61 | 0.00 | 0.00 | 0.00 | 0.00 | 0.00 | 0.61 | Virtual elimination |
| 0.7 | 0.3 | Permanent | Winter | 0 | 0.00 | 0.34 | 0.03 | 0.00 | 0.00 | 0.00 | 0.00 | 0.34 | Virtual elimination |
| 0.3 | 0 | Permanent | Winter | 0 | 0.00 | 0.61 | 0.00 | 0.00 | 0.00 | 0.00 | 0.00 | 0.61 | Virtual elimination |
| 0 | 0 | Permanent | Winter | 0 | 0.00 | 0.61 | 0.00 | 0.00 | 0.00 | 0.00 | 0.00 | 0.61 | Virtual elimination |
| 0.7 | 0 | 40 | Spring | 0 | 0.00 | 0.63 | 0.57 | 0.17 | 0.09 | 0.07 | 0.07 | 0.63 | Annual |
| 0.7 | 0.3 | 40 | Spring | 0 | 0.00 | 0.34 | 0.40 | 0.14 | 0.08 | 0.07 | 0.07 | 0.40 | Annual |
| 0.3 | 0 | 40 | Spring | 0 | 0.00 | 0.63 | 0.57 | 0.17 | 0.09 | 0.07 | 0.07 | 0.63 | Annual |
| 0 | 0 | 40 | Spring | 0 | 0.00 | 0.63 | 0.57 | 0.17 | 0.09 | 0.07 | 0.07 | 0.63 | Annual |
| 0.7 | 0 | 104 | Spring | 0 | 0.00 | 0.62 | 0.55 | 0.03 | 0.16 | 0.08 | 0.04 | 0.62 | Annual |
| 0.7 | 0.3 | 104 | Spring | 0 | 0.00 | 0.34 | 0.39 | 0.12 | 0.07 | 0.06 | 0.04 | 0.39 | Annual |
| 0.3 | 0 | 104 | Spring | 0 | 0.00 | 0.62 | 0.55 | 0.03 | 0.16 | 0.08 | 0.04 | 0.62 | Annual |
| 0 | 0 | 104 | Spring | 0 | 0.00 | 0.62 | 0.55 | 0.03 | 0.16 | 0.08 | 0.04 | 0.62 | Annual |
| 0.7 | 0 | Permanent | Spring | 0 | 0.00 | 0.61 | 0.54 | 0.00 | 0.00 | 0.00 | 0.00 | 0.61 | Virtual elimination |
| 0.7 | 0.3 | Permanent | Spring | 0 | 0.00 | 0.33 | 0.38 | 0.00 | 0.00 | 0.00 | 0.00 | 0.38 | Virtual elimination |
| 0.3 | 0 | Permanent | Spring | 0 | 0.00 | 0.61 | 0.54 | 0.00 | 0.00 | 0.00 | 0.00 | 0.61 | Virtual elimination |
| 0 | 0 | Permanent | Spring | 0 | 0.00 | 0.61 | 0.54 | 0.00 | 0.00 | 0.00 | 0.00 | 0.61 | Virtual elimination |
| 0.7 | 0 | 40 | Summer | 0 | 0.00 | 0.00 | 0.63 | 0.17 | 0.09 | 0.08 | 0.07 | 0.63 | Annual |
| 0.7 | 0.3 | 40 | Summer | 0 | 0.00 | 0.00 | 0.45 | 0.15 | 0.09 | 0.07 | 0.08 | 0.45 | Annual |
| 0.3 | 0 | 40 | Summer | 0 | 0.00 | 0.00 | 0.63 | 0.17 | 0.09 | 0.08 | 0.07 | 0.63 | Annual |
| 0 | 0 | 40 | Summer | 0 | 0.00 | 0.00 | 0.63 | 0.17 | 0.09 | 0.08 | 0.07 | 0.63 | Annual |
| 0.7 | 0 | 104 | Summer | 0 | 0.00 | 0.00 | 0.62 | 0.00 | 0.16 | 0.07 | 0.08 | 0.62 | Annual |
| 0.7 | 0.3 | 104 | Summer | 0 | 0.00 | 0.00 | 0.44 | 0.04 | 0.13 | 0.04 | 0.03 | 0.44 | Annual |
| 0.3 | 0 | 104 | Summer | 0 | 0.00 | 0.00 | 0.62 | 0.00 | 0.16 | 0.07 | 0.08 | 0.62 | Annual |
| 0 | 0 | 104 | Summer | 0 | 0.00 | 0.00 | 0.62 | 0.00 | 0.16 | 0.07 | 0.08 | 0.62 | Annual |
| 0.7 | 0 | Permanent | Summer | 0 | 0.00 | 0.00 | 0.61 | 0.00 | 0.00 | 0.00 | 0.00 | 0.61 | Virtual elimination |
| 0.7 | 0.3 | Permanent | Summer | 0 | 0.00 | 0.00 | 0.43 | 0.00 | 0.00 | 0.00 | 0.00 | 0.43 | Virtual elimination |
| 0.3 | 0 | Permanent | Summer | 0 | 0.00 | 0.00 | 0.61 | 0.00 | 0.00 | 0.00 | 0.00 | 0.61 | Virtual elimination |
| 0 | 0 | Permanent | Summer | 0 | 0.00 | 0.00 | 0.61 | 0.00 | 0.00 | 0.00 | 0.00 | 0.61 | Virtual elimination |
| 0.7 | 0 | 40 | Autumn | 0 | 0.00 | 0.00 | 0.63 | 0.17 | 0.09 | 0.08 | 0.07 | 0.63 | Annual |
| 0.7 | 0.3 | 40 | Autumn | 0 | 0.00 | 0.00 | 0.47 | 0.15 | 0.09 | 0.08 | 0.08 | 0.47 | Annual |
| 0.3 | 0 | 40 | Autumn | 0 | 0.00 | 0.00 | 0.63 | 0.17 | 0.09 | 0.08 | 0.07 | 0.63 | Annual |
| 0 | 0 | 40 | Autumn | 0 | 0.00 | 0.00 | 0.63 | 0.17 | 0.09 | 0.08 | 0.07 | 0.63 | Annual |
| 0.7 | 0 | 104 | Autumn | 0 | 0.00 | 0.00 | 0.62 | 0.00 | 0.16 | 0.03 | 0.08 | 0.62 | Biennial |
| 0.7 | 0.3 | 104 | Autumn | 0 | 0.00 | 0.00 | 0.46 | 0.00 | 0.13 | 0.05 | 0.05 | 0.46 | Biennial |
| 0.3 | 0 | 104 | Autumn | 0 | 0.00 | 0.00 | 0.62 | 0.00 | 0.16 | 0.03 | 0.08 | 0.62 | Biennial |
| 0 | 0 | 104 | Autumn | 0 | 0.00 | 0.00 | 0.62 | 0.00 | 0.16 | 0.03 | 0.08 | 0.62 | Biennial |
| 0.7 | 0 | Permanent | Autumn | 0 | 0.00 | 0.00 | 0.61 | 0.00 | 0.00 | 0.00 | 0.00 | 0.61 | Virtual elimination |
| 0.7 | 0.3 | Permanent | Autumn | 0 | 0.00 | 0.00 | 0.46 | 0.00 | 0.00 | 0.00 | 0.00 | 0.46 | Virtual elimination |
| 0.3 | 0 | Permanent | Autumn | 0 | 0.00 | 0.00 | 0.61 | 0.00 | 0.00 | 0.00 | 0.00 | 0.61 | Virtual elimination |
| 0 | 0 | Permanent | Autumn | 0 | 0.00 | 0.00 | 0.61 | 0.00 | 0.00 | 0.00 | 0.00 | 0.61 | Virtual elimination |

**Table S7.** Parameter values for the two-strain SEIR transmission model representing HCoV-OC43 (strain 1) and HCoV-HKU1 (strain 2).

| Parameter | Range for LHS | Interpretation | Units | Best value (95% CI) |
| --- | --- | --- | --- | --- |
| $a$ | [0, 2] | Seasonal $R_0$ amplitude | none | 0.66 (0.53, 0.98) |
| $b$ | [1, 2] | Seasonal $R_0$ baseline | none | 1.4 (1.3, 1.5) |
| $\phi$ | [-8, 8] | Seasonal $R_0$ shift | weeks | -3.8 (-5.8, -1.7) |
| $\sigma_1$ | [25, 100] | Waning immunity period, strain 1 | weeks | 40 (31, 52) |
| $\sigma_2$ | [25, 100] | Waning immunity period, strain 2 | weeks | 38 (31, 47) |
| $\chi_{12}$ | [0, 1] | Cross immunity, strain 1 against strain 2 | none | 0.74 (0.67, 0.78) |
| $\chi_{21}$ | [0, 1] | Cross immunity, strain 2 against strain 1 | none | 0.50 (0.10, 0.65) |
| $1/\nu$ | [3, 14] | Incubation period | days | 5.0 (3.4, 6.0) |
| $1/\gamma$ | [3, 14] | Infectious period | days | 4.9 (3.3, 5.9) |
| $1/\mu$ | 80 (fixed) | Average lifespan | years | 80 (fixed) |

**Figure S1.**
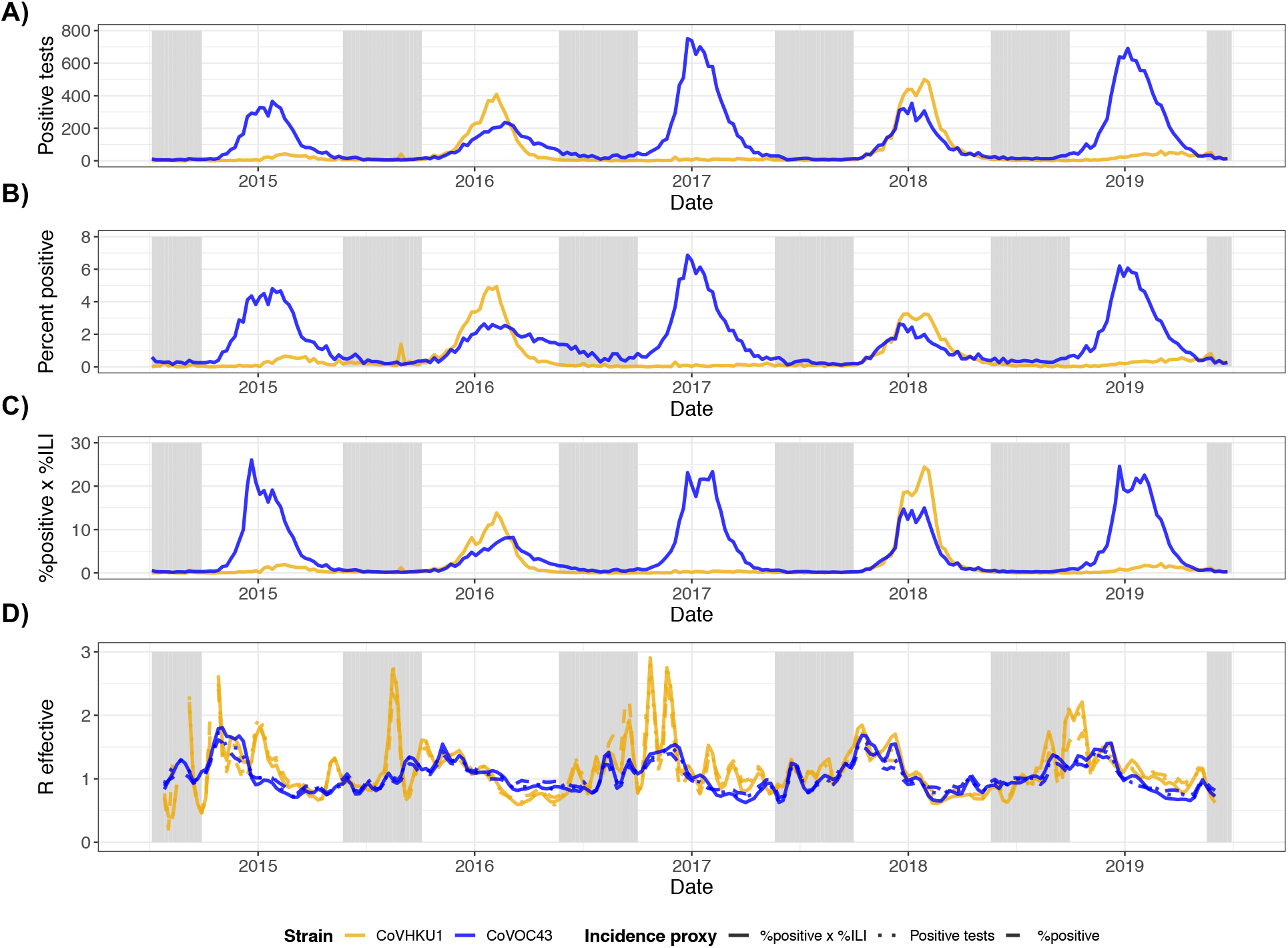
Incidence proxies and effective reproduction numbers for HCoV-HKU1 (gold) and HCoV-OC43 (blue). (A) Weekly number of positive tests from NREVSS. (B) Weekly percent positive laboratory tests. (C) Weekly percent positive laboratory tests multiplied by percent of clinic visits for ILI. (D) Comparison of effective reproduction numbers by incidence measure using SARS serial interval. R effective estimates for HCoV-HKU1 that were greater than 3 (all occurring at the end of 2014) are not shown.

**Figure S2.**
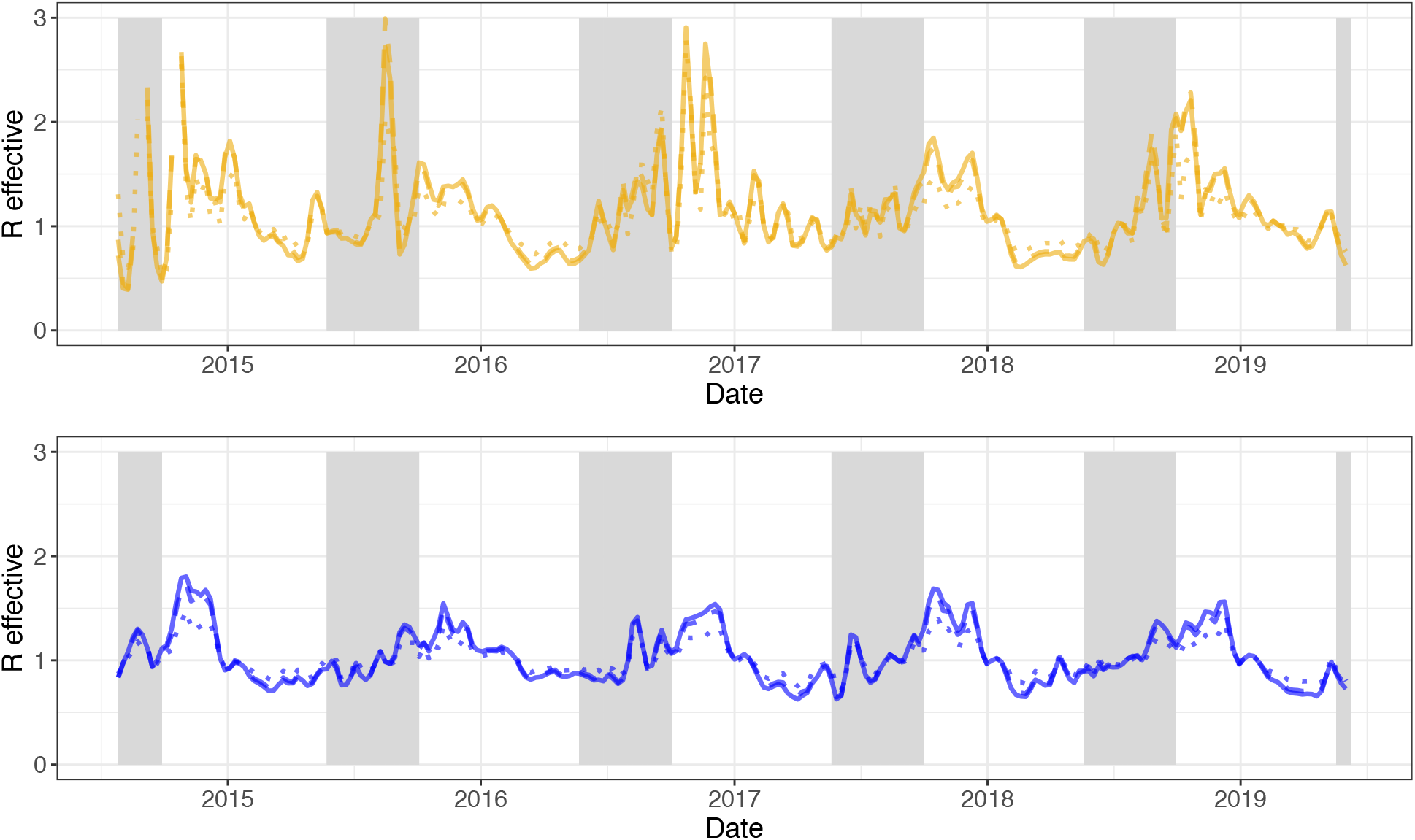
Estimates of weekly effective reproduction numbers for HCoV-HKU1 (top) and HCoV-OC43 (bottom) based on different serial interval distributions. Serial interval distributions were defined as follows - SARS (solid): Weibull distribution with mean of 8.4 days and s.d. of 3.8 days (shape=2.35, scale=9.48); Li2020 (dashed): Gamma distribution with mean of 7.5 days and s.d. of 3.4 days (shape=4.87, scale=1.54); Nishiura2020 (dotted): Weibull distribution with mean of 4.8 days and s.d. of 2.3 days (shape=2.20, scale=5.42). Sections shaded in gray are out-of-season (epidemiological weeks 21-39). R effective estimates for HCoV-HKU1 that were greater than 3 (all occurring at the end of 2014) are not shown.

**Figure S3.**
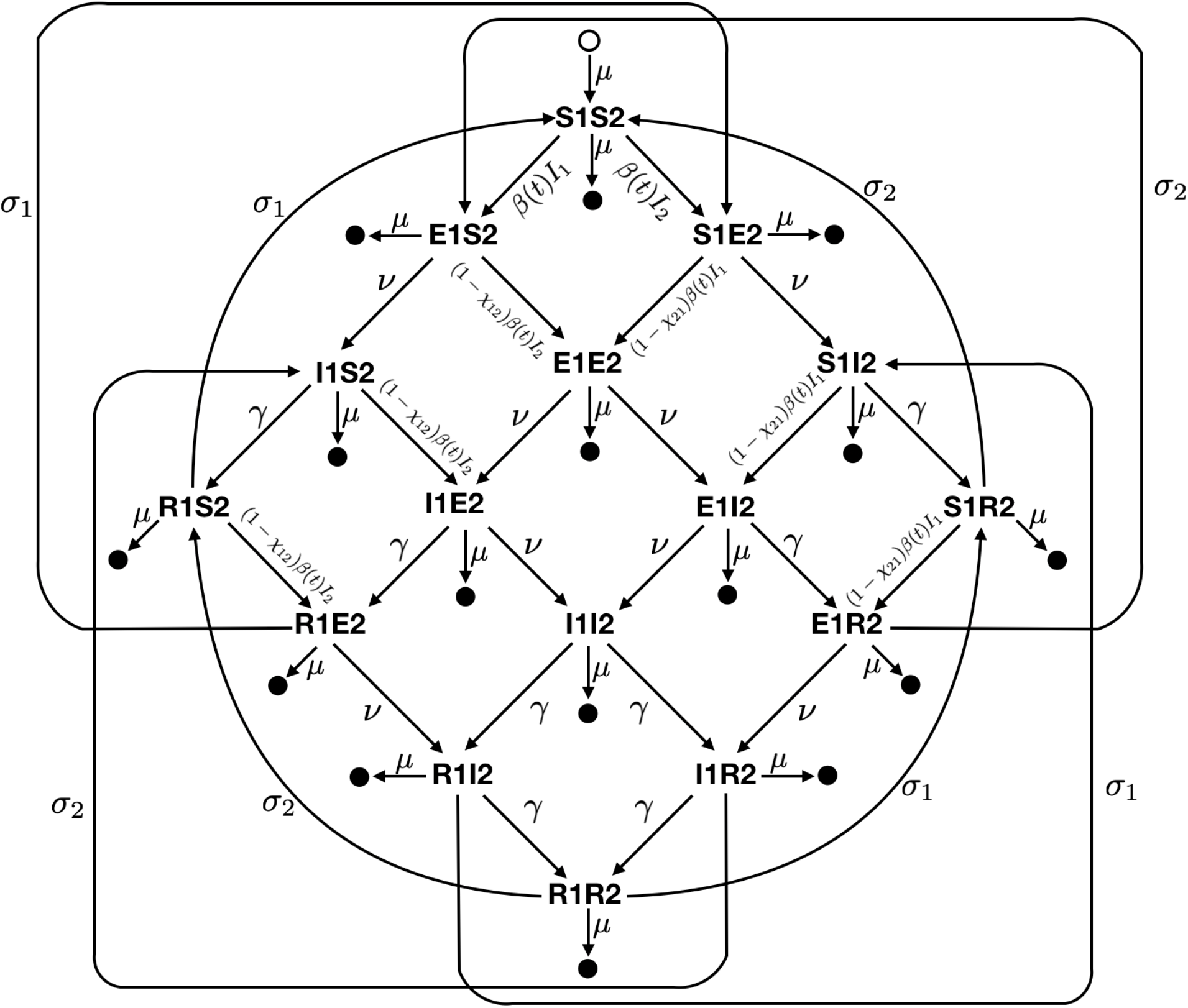
Diagram of the two-strain compartmental SEIR model used to describe the transmission of HCoV-OC43 and HCoV-HKU1 in the United States. Epidemiological compartments are represented by the bold letter-number pairs, such that an individual in compartment S1S2 is susceptible to both strains, while a person in compartment I1E2 is infectious with strain 1 and has been exposed to strain 2. Filled circles represent death and the open circle represents births. Transition rates are given next to the arrows between compartments. Best-fit parameter values are listed in **Table S7**.

